# The co-occurrence of genetic variants in the *TYR* and *OCA2* genes confers susceptibility to albinism

**DOI:** 10.1101/2023.01.19.23284597

**Authors:** David J. Green, Vincent Michaud, Eulalie Lasseaux, Claudio Plaisant, UK Biobank Eye and Vision Consortium, Tomas Fitzgerald, Ewan Birney, Graeme C. Black, Benoît Arveiler, Panagiotis I. Sergouniotis

## Abstract

Although rare genetic conditions are mostly caused by DNA sequence alterations that functionally disrupt individual genes, large-scale studies using genome sequencing have started to unmask additional complexity. Understanding how combinations of variants in different genes shape human phenotypes is expected to provide important insights into the clinical and genetic heterogeneity of rare disorders. We used albinism, an archetypal rare condition associated with hypopigmentation, as an exemplar for the study of genetic interactions. We analysed data from the Genomics England 100,000 Genomes Project alongside a cohort of 1,120 individuals with albinism, and investigated the effect of dual heterozygosity for the combination of two established albinism-related variants: *TYR*:c.1205G>A (p.Arg402Gln) [rs1126809] and *OCA2*:c.1327G>A (p.Val443Ile) [rs74653330]. As each of these changes alone is insufficient to cause disease when present in the heterozygous state, we sought evidence of synergistic effects. We found that, when both variants are present, the probability of receiving a diagnosis of albinism is significantly increased (odds ratio 12.8; 95% confidence interval 6.0 – 24.7; p-value 2.1 x 10^-8^). Further analyses in an independent cohort, the UK Biobank, supported this finding and highlighted that heterozygosity for the *TYR*:c.1205G>A and *OCA2*:c.1327G>A variant combination is associated with statistically significant alterations in visual acuity and central retinal thickness (traits that are considered albinism endophenotypes). The approach discussed in this report opens up new avenues for the investigation of oligogenic patterns in apparently Mendelian disorders.

## INTRODUCTION

An individual affected by a condition that appears to segregate as a Mendelian trait, would be expected to carry large-effect genetic variation in a single gene/locus. However, for a significant proportion of affected probands, single-gene-centric genomic analyses fail to identify a clear genetic diagnosis.^1–6^ Although this diagnostic and knowledge gap is, to a degree, due to imperfect phenotyping, sequencing or annotation, other factors may be contributing. There is for example growing evidence supporting the hypothesis that many apparently Mendelian phenotypes are caused by the interaction of multiple variants in more than one locus.^7–11^ Uncovering such genetic interactions has the potential to provide insights not only into disease mechanisms and diagnostics but also into phenomena like incomplete penetrance and variable expressivity.^12–14^

There are many patterns through which genetic changes in more than one gene can synergistically shape a phenotype. A simple scenario, would involve two heterozygous variants in two distinct loci cooperating to mediate disease (when each individual variant in isolation fails to explain the clinical presentation). Only a small number of cases with this specific form of digenic inheritance have been described^15–17^, the majority detected through qualitative approaches involving variant segregation analyses in a small number of families. In these cases, interaction-based models have generally been used to provide a post hoc explanation of the observed phenomena. By contrast, we describe a quantitative, case-control approach that allows obtaining statistical evidence of digenic patterns. We focused on specific genotypes in the *TYR* and *OCA2* genes; biallelic variants in each of these two genes are associated with albinism, a clinically and genetically heterogeneous group of conditions characterised by reduced levels of melanin pigment. Key features of albinism include visual abnormalities and under-development of the central retina (*i.e.* the fovea); skin and/or hair hypopigmentation may also be present.^18^ Building on recent work^19–22^, we aimed to advance our understanding of the genetic complexity of this rare disorder.

## RESULTS & DISCUSSION

A cohort of 1,015 people with albinism underwent testing of ≤19 albinism-related genes; these individuals were not known to be related and had predominantly European-like ancestries (Supplementary Data 1). A further 105 probands with albinism were identified in the Genomics England 100,000 Genomes Project dataset.^23^ A “control” cohort of 29,451 unrelated individuals that had no recorded diagnosis or features of albinism was also identified in this resource (Fig. 1, Supplementary Table 1, Methods).

**Figure 1.**
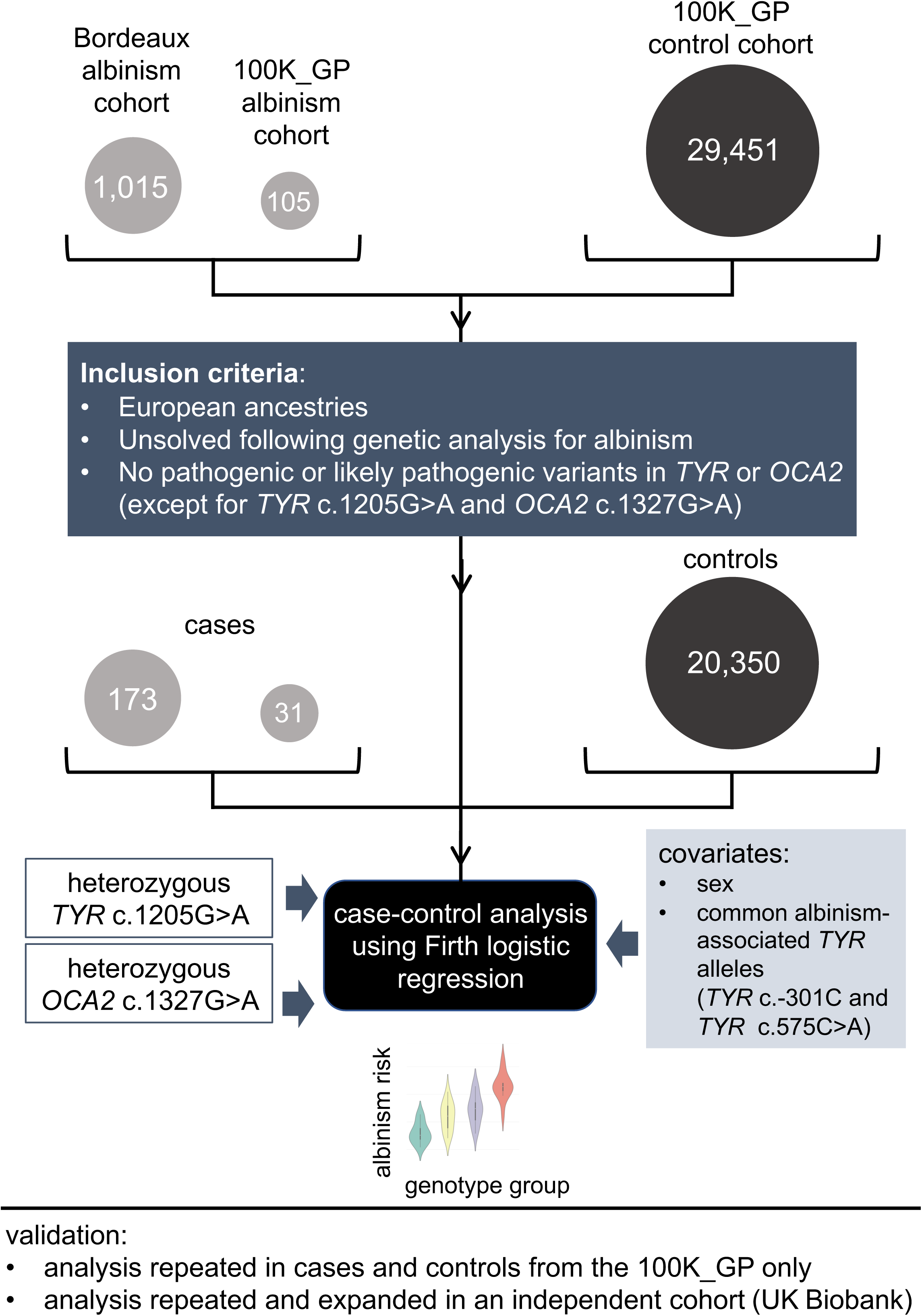
Outline of the case-control study design. A case-control analysis was performed to gain insights into the role of genotypes involving dual heterozygosity for *TYR*:c.1205G>A (p.Arg402Gln) [rs1126809] and *OCA2*:c.1327G>A (p.Val443Ile) [rs74653330] in albinism. These two missense changes were selected as they are the commonest albinism-related variants in European-like populations. The majority of participants in the “case” cohort (1,015/1,120) were identified through the database of the University Hospital of Bordeaux Molecular Genetics Laboratory, France. All these probands had at least one key ocular feature of albinism, *i.e.* nystagmus or prominent foveal hypoplasia (see Methods). The remaining 105/1,120 cases were identified through the Genomics England 100,000 Genomes Project dataset and had a diagnosis of albinism or partial/ocular albinism. The “control” cohort included 29,451 unrelated individuals from the Genomics England 100,000 Genomes Project dataset, none of whom had a recorded diagnosis of albinism. To reduce the likelihood of obtaining spurious signals due to population stratification effects or due to the presence of albinism-related variants other than the two studied changes, we focused only on individuals who: (i) were projected to have European-like ancestries; (ii) did not have a genotype in keeping with a molecular diagnosis of albinism; (iii) were not heterozygous for a pathogenic or a likely pathogenic variant in *TYR* or *OCA2* (excluding *TYR*:c.1205G>A and *OCA2*:c.1327G>A). The following two covariates were used: sex and number of common albinism-associated alleles, *i.e.* DNA sequence alterations in the genomic locations corresponding to *TYR*:c.−301C>T [rs4547091] and *TYR*:c.575C>A (p.Ser192Tyr) [rs1042602]. These two variants have been previously shown to modify the effect of *TYR*:c.1205G>A which is a common missense change that can act as a “hypomorphic” variant.^19–21^ To assess the robustness of the findings we performed additional analyses in subsets of the cohort. Validation studies in an independent cohort (UK Biobank) were also conducted. 100K_GP, Genomics England 100,000 Genomes Project; het, heterozygous; UKB, UK Biobank. *TYR* and *OCA2* variant numbering was based on the transcripts with the following identifiers: NM_000372.5/ENST00000263321.6. and NM_000275.3/ENST00000354638.8.

Searching for genetic elements underlying digenic traits may be carried out at the level of genes, genotypes or DNA variants. It is noted that a given pair of DNA variants comprises up to nine pairs of genotypes while a given gene may contain a large number of variants. Each of these search strategies has inherent advantages and pitfalls.^24^ We opted for a genotype-based approach that offers a high level of precision and is more likely to provide insights that can be used for clinical genome interpretation.

Our genotype-based analysis focused on two prevalent albinism-related changes, *TYR*:c.1205G>A (p.Arg402Gln) [rs1126809] and *OCA2*:c.1327G>A (p.Val443Ile) [rs74653330]. We hypothesised that when these two missense variants are both present in the heterozygous state (“dual heterozygosity”), their interaction is driving the pathology observed in albinism. Notably, both these changes have been shown to reduce the activity of their respective protein products *in vitro*.^25–29^ Further, previous studies have pointed to a functional interaction between the TYR and OCA2 proteins: the latter has a role in the pH regulation of the melanosome which, in turn, can affect the catalytic activity of the former (*i.e.* tyrosinase, the rate-limiting enzyme for the synthesis of melanin in the melanosomes of retinal pigment epithelia and melanocytes).^18,30^ Both *TYR*:c.1205G>A and *OCA2*:c.1327G>A have been implicated in albinism when *in trans* with pathogenic changes in the same gene.^21,31^ Also, multiple associations have been recorded for these two variants including with skin/hair pigmentation and skin cancer.^32^ It is noted that these two changes were chosen as they are the two commonest albinism-related variants in populations of European-like ancestries, a feature that can help mitigate potential issues with statistical power. It is also highlighted that the minor allele frequencies of the *TYR*:c.1205G>A and *OCA2*:c.1327G>A changes in European populations are 27% and 0.3% respectively (Non-Finnish European subset of the Genome Aggregation Database [gnomAD] v2.1.1).^33^ The particularly common *TYR*:c.1205G>A change has been associated with incomplete penetrance (and variable expressivity), and previous studies have pointed to the potential for other *TYR* variants to modify its effect.^19–21,25,34–36^

We used Firth regression analysis^37,38^ to study how dual heterozygosity for *TYR*:c.1205G>A and *OCA2*:c.1327G>A affects the probability of receiving a diagnosis of albinism (“risk of albinism”). This logistic regression approach has been designed to handle small, imbalanced datasets (which are common in studies of rare conditions) and allows for adjustment of key covariates (which is not possible in contingency table methods) (Fig.1). To increase the genetic similarity^39^ between the compared cohorts (cases and controls) and to reduce the impact of albinism-related variants other than the two studied changes (*TYR*:c.1205G>A and *OCA2*:c.1327G>A), we excluded from our analysis individuals that: (i) did not have European-like ancestries, (ii) were found to have genotypes in keeping with a molecular diagnosis of albinism, (iii) carried pathogenic or likely pathogenic variants in the *TYR* or *OCA2* gene (with the exception of *TYR*:c.1205G>A and *OCA2*:c.1327G>A). The resulting case and control cohorts included 204 and 20,350 individuals respectively. These were used to perform case-control comparisons between the following genotype groups:

- *group A* (reference group): homozygous for *TYR*:c.1205= and *OCA2*:c.1327=
- *group B* (single *TYR* heterozygote group): heterozygous for *TYR*:c.1205G>A and homozygous for *OCA2*:c.1327=.
- *group C* (single *OCA2* heterozygote group): homozygous for *TYR*:c.1205G= and heterozygous for *OCA2*:c.1327G>A.
- *group D* (dual heterozygote group): heterozygous for the *TYR*:c.1205G>A and *OCA2*:c.1327G>A variant combination.

These groups correspond to different levels of dysfunction of the TYR and OCA2 molecules and the findings of the regression analysis revealed a synergistic potentiation of the changes at the two loci (Fig.2, Supplementary Fig. 1a and Supplementary Table 2). The point estimate for the odds of receiving a diagnosis of albinism in individuals who are heterozygous for the *TYR*:c.1205G>A and *OCA2*:c.1327G>A variant combination was 12.8 (the 95% confidence interval was 6.0 – 24.7 and the p-value was 2.1 x 10^-8^).

**Figure 2.**
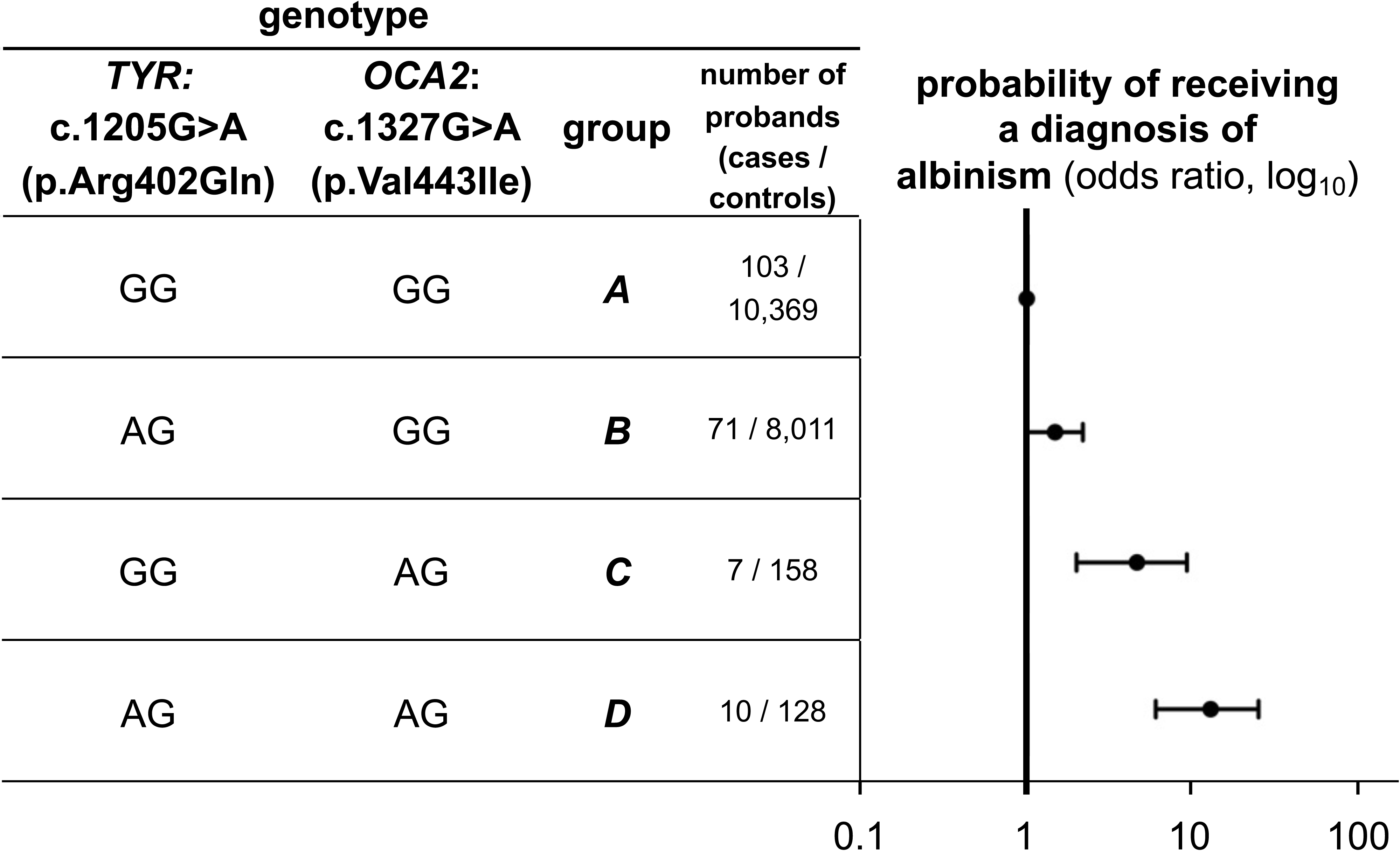
The combination of the *TYR*:c.1205G>A (p.Arg402Gln) and *OCA2*:c.1327G>A (p.Val443Ile) variants in a dual heterozygous state confers susceptibility to albinism. The probability of receiving a diagnosis of albinism (*i.e.* the risk of albinism) was calculated for the following genotype groups:

- ***group A*** (reference group): homozygous for *TYR*:c.1205= and *OCA2*:c.1327=
- ***group B*** (single *TYR* heterozygote group): heterozygous for *TYR*:c.1205G>A and homozygous for *OCA2*:c.1327=
- ***group C*** (single *OCA2* heterozygote group): homozygous for *TYR*:c.1205G= and heterozygous for *OCA2*:c.1327G>A
- ***group D*** (dual heterozygote group): heterozygous for the *TYR*:c.1205G>A and *OCA2*:c.1327G>A variant combination. A log_10_ scale is used. The circle in the middle of each horizontal line (95% confidence interval) represents the point estimate of each odds ratio. Group A was used as the reference group to which all other groups were compared using Firth regression analysis (*i.e.* the odds ratio for this group was fixed at 1). Further information including numerical data can be found in Supplementary Table 2.

To increase confidence in these observations, we performed additional analyses in an independent cohort, the UK Biobank.^40^ We found that UK Biobank volunteers who were heterozygous for the *TYR*:c.1205G>A and *OCA2*:c.1327G>A variant combination had not only a higher chance of receiving a diagnosis of albinism (OR>6.9, p-value=0.0004) but also had, on average, slightly worse visual acuity and marginally thicker central retina (the respective Kruskal-Wallis p-values were 1.4 x 10^-3^ and 8.2 x 10^-4^; Fig. 3 and Supplementary Tables 3-6). It is noted that visual acuity and central retinal thickness are quantitative endophenotypes of albinism and these findings suggest that the studied *TYR*/*OCA2* variant combination is both contributing to the genetic architecture of albinism and having a role in the development of the visual system in individuals from the general population.

**Figure 3.**
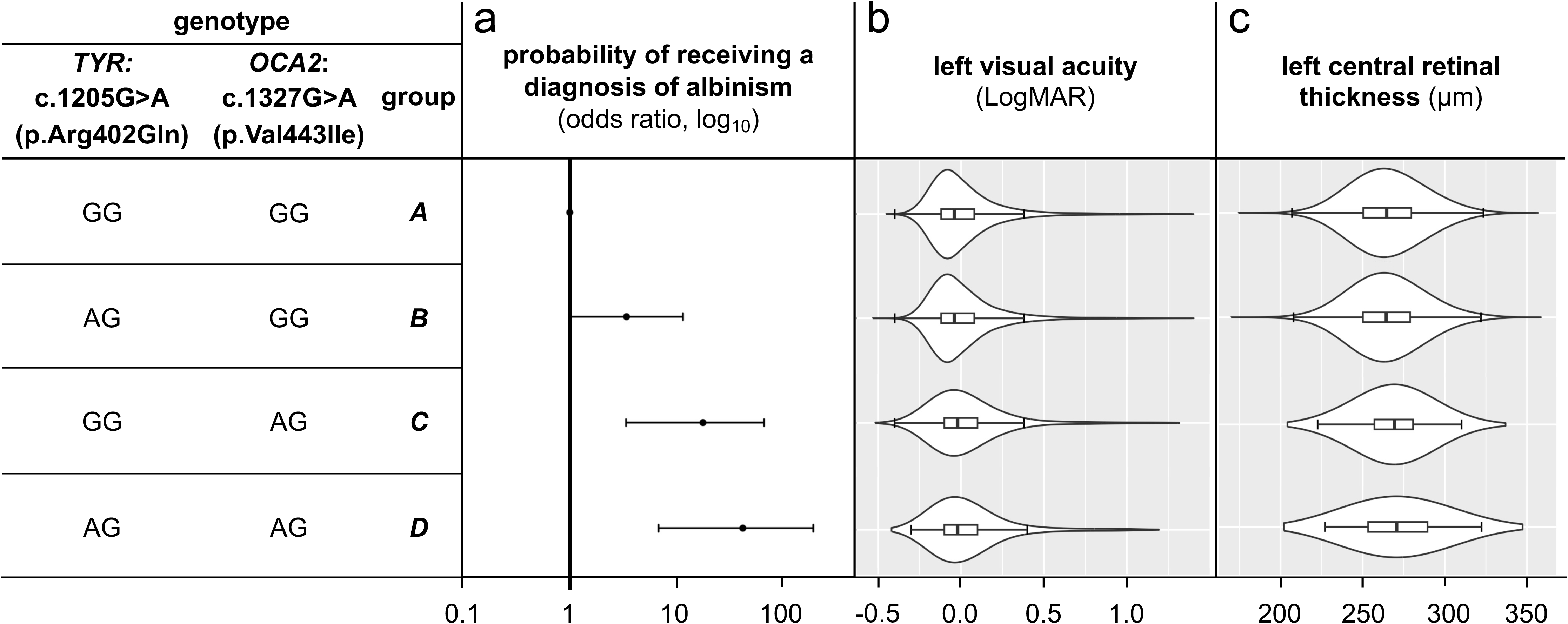
The combination of the *TYR*:c.1205G>A (p.Arg402Gln) and *OCA2*:c.1327G>A (p.Val443Ile) variants in a dual heterozygous state is associated with (a) a higher probability of receiving an albinism diagnosis (*i.e.* a higher risk of albinism), (b) lower visual acuity and (c) increased central retinal thickness in UK Biobank participants. The following genotype groups were studied:

- ***group A*** (reference group): homozygous for *TYR*:c.1205= and *OCA2*:c.1327=
- ***group B*** (single *TYR* heterozygote group): heterozygous for *TYR*:c.1205G>A and homozygous for *OCA2*:c.1327=
- ***group C*** (single *OCA2* heterozygote group): homozygous for *TYR*:c.1205G= and heterozygous for *OCA2*:c.1327G>A
- ***group D*** (dual heterozygote group): heterozygous for the *TYR*:c.1205G>A and *OCA2*:c.1327G>A variant combination. It is noted that individuals with albinism tend to have reduced visual acuity (*i.e.* higher LogMAR value than 0.2) and increased central retinal thickness (due to underdevelopment of the fovea). More details on the utilised approach can be found in the Methods and in Supplementary Fig. 2 and 3. Further information including numerical data can be found in Supplementary Tables 3-6.

We provide evidence that the *TYR*:c.1205G>A and *OCA2*:c.1327G>A changes appear to have a joint effect when each is encountered in the heterozygous state. However, the interpretation of this variant combination in clinical diagnostic settings should be approached cautiously. Although the detected effect size (odds ratio) is significant, it would be more in keeping with a predisposing^41^ rather than a pathogenic^42^ genotype. Further, while it is tempting to view our observations as an example of a digenic pattern, this would have reductionist connotations; interactions are ubiquitous in nature and we expect that increased sample sizes will elucidate longer “chains” of interconnected genetic variation. It is thus plausible that the effect size of the studied genotype will increase when further genetic changes that impact on tyrosinase function or melanosome pH are added to the model. Indeed, observations in the presented data already point to this possibility (Supplementary Fig. 3; Supplementary Data 2 and 3).

One limitation of this study is our inability to match stringently the albinism cases with the unaffected controls, especially in terms of genetic background.^43^ Although recent ancestry was considered, our analysis was imperfect as it was not possible to reliably assign genetic ancestry to most albinism cases. We used a combination of orthogonal approaches to evaluate the robustness and generalisability of our findings. First, we used 35 presumed neutral single-nucleotide variants to calculate the genomic inflation factor lambda (λGC)^44,45^; the λ_median_ was found to be 1.04, in keeping with limited confounding by ancestry (Supplementary Table 7). Subsequently, we performed a targeted secondary analysis using only Genomics England 100,000 Genomes Project data which allowed us to study case and control groups of high genetic similarity. The results supported our key findings and increased confidence in the validity of the detected associations (Supplementary Fig. 1b and Supplementary Table 8). Another potential caveat is that the detected signal in dual heterozygotes for *TYR*:c.1205G>A and *OCA2*:c.1327G>A could, to a degree, be driven by undetected pathogenic variation in either *TYR* or *OCA2*. This cannot be excluded despite the fact that medical-grade genetic analyses were undertaken. It can be argued however that significant confounding is unlikely especially given that the effect of dual heterozygosity for *TYR*:c.1205G>A and *OCA2*:c.1327G>A appears to be consistently greater than that of genotypes involving heterozygosity for either *TYR*:c.1205G>A or *OCA2*:c.1327G>A alone (Supplementary Tables 2, 3, 8 and 9). Nonetheless, statistically significant results were also obtained for individuals carrying the *OCA2:*c.1327G>A change in heterozygous state (without TYR:c.1205G>A) and future studies will provide further insights into the extent to which this change alone drives the observed signal.

In conclusion, we have shown that dual heterozygosity for a *TYR* and an *OCA2* variant confers susceptibility to albinism. Shedding light into this and other variant interactions is expected to increase our understanding of the pathways and cellular processes affected in albinism. Notably, the concepts discussed herein are likely to be relevant to other rare disorders, and systems-based approaches using biological modules are expected to narrow the molecular diagnostic gap. Future work will focus on increasing sample sizes, embracing more diverse populations and searching for high-order interactions by considering more than two variants at a time.

## METHODS

### Cohort characteristics and genotyping

#### University Hospital of Bordeaux albinism cohort

Individuals with albinism were identified through the database of the University Hospital of Bordeaux Molecular Genetics Laboratory, France. This is a national reference laboratory that has been performing genetic testing for albinism since 2003 and has been receiving samples from individuals predominantly based in France (or French-administered overseas territories).

Information on the dermatological and ophthalmological phenotypes was available for all affected individuals. The relevant clinical data were assessed independently by two clinicians with experience in the field of albinism and only cases with a consensus clinical impression of albinism were included. It is noted that and each of these cases had at least one of the key ocular features of albinism, *i.e.* infantile nystagmus or absence of a foveal pit (prominent foveal hypoplasia) (Supplementary Table 10). No pre-screening based on genotype was undertaken other than selecting individuals in whom position c.-301 of *TYR* was sequenced. Only individuals who were not knowingly related were included.

Genetic testing, bioinformatic analyses, and variant interpretation were performed as previously described.^20,21^ Briefly, most participants had gene-panel testing of ≤19 genes associated with albinism (*C10ORF11*, *GPR143*, *HPS 1* to 10, *LYST, OCA2*, *SLC38A8, SLC24A5*, *SLC45A2*, *TYR*, *TYRP1*) using IonTorrent platforms. High-resolution array-CGH (comparative genomic hybridization) was used to detect copy number variants in these genes. All genetic changes of interest were confirmed with an alternative method (e.g. Sanger sequencing or quantitative PCR). Clinical interpretation of variants was performed using criteria consistent with the 2015 American College of Medical Genetics and Genomics (ACMG) best practice guidelines.^42^ It is noted that genetic findings in this cohort have been partly reported in previous publications from our group^19,22^ and that comprehensive genomic testing (including single-nucleotide variant and copy number variant analysis involving the aforementioned 19 albinism-related genes) was performed in all cases that remained unsolved.

Due to the limited number of genomic loci screened in this cohort, it was not possible to reliably assess genetic ancestry and to “objectively” assign individuals to genetic ancestry groups. Attempting to mitigate this, we processed available data on self-identified ethnicity that were collected through questionnaires. Responses were inspected and stratification into five broad continental groups (European, African, Admixed American, East Asian, South Asian) was performed.

#### Genomics England 100,000 Genomes Project cohort

Clinical and genomic data from the Genomics England 100,000 Genomes Project were accessed through a secure Research Environment that is available to registered users. This dataset was collected as part of a national genome sequencing initiative.^46^ Enrolment was coordinated by Genomics England Limited, and participants were recruited mainly at National Health Service (NHS) Hospitals in the UK.^23^ Clinical information was recorded in Human Phenotype Ontology (HPO)^47^ terms and International Classification of Diseases (ICD) codes. Genome sequencing was performed in DNA samples from ≥78,195 individuals using Illumina HiSeq X systems (150 base-pair paired-end format). Reads were aligned using the iSAAC Aligner v03.16.02.19 and small variants were called using Starling v2.4.7.^48^ Structural variants and long indel (>50bp) calling was performed with Manta v0.28.0, and copy number variants (CNVs) were called with Canvas v1.3.1.^49,50^ Aggregation of single-sample gVCFs was performed using the Illumina software gVCF genotyper v2019.02.29; normalisation/decomposition was implemented by vt v0.57721^51^. The multi-sample VCF was then split into 1,371 roughly equal chunks to allow faster processing and the loci of interest were queried using bcftools v1.9^52^ (see https://research-help.genomicsengland.co.uk/display/GERE/ for further information). Only variants that passed standard quality control criteria were processed (including the criteria discussed in https://re-docs.genomicsengland.co.uk/site_qc/ and criteria analogous to those outlined in https://re-docs.genomicsengland.co.uk/somatic_sv/#note-on-variant-calling). In addition, we filtered out genotypes with: genotype score <20; read depth <10; allele balance <0.2 and >0.8 for heterozygotes; allele balance >0.1 or <0.9 for homozygotes (reference and alternate, respectively). Genomic annotation was performed using Ensembl VEP^53^. Variants with a “total” minor allele frequency (MAF) <1% in the Genome Aggregation Database (gnomAD v2.1.1)^33^ and a “disease-causing” (DM) label in the Human Gene Mutation Database (HGMD) v2021.2^54^ were flagged (and are hitherto referred to as “HGMD-listed variants”). In depth manual curation of all detected rare (*i.e.* MAF < 1%) variants in *TYR* and *OCA2* was undertaken and these changes were classified according to the 2015 American College of Medical Genetics and Genomics (ACMG) best practice guidelines.^42^ Ancestry inference was performed in the Genomics England 100,000 Genomes Project cohort using principal component analysis. Data from the 1000 genomes project (phase 3) dataset were used and five broad super-populations were projected (European, African, Admixed American, East Asian, South Asian) (further information on this can be found online at https://research-help.genomicsengland.co.uk/display/GERE/Ancestry+inference).

We focused on a pre-determined subset of the Genomics England 100,000 Genomes Project dataset that includes only unrelated probands (n=29,556). Of these, 105 had a diagnosis of albinism, *i.e.* the ICD-10 term “Albinism” [E70.3] and/or the HPO terms “Albinism” [HP:0001022], “Partial albinism” [HP:0007443] or “Ocular albinism” [HP:0001107] were assigned (Supplementary Table 10). Together with the University Hospital of Bordeaux cases, these 105 probands formed the “case” cohort. The remaining 29,451 probands had no recorded diagnosis or phenotypic features of albinism and formed the “control” cohort. We note that comprehensive ophthalmic phenotyping was not routinely undertaken in Genomics England 100,000 Genomes Project participants. We cannot therefore be certain that a small number of individuals with mild/subclinical forms of albinism are not included in the control cohort.

### Case-control analysis to estimate the probability of an albinism diagnosis

The effect of dual heterozygosity for *TYR:*c.1205G>A (p.Arg402Gln) [rs1126809] and *OCA2:*c.1327G>A (p.Val443Ile) [rs74653330] on the probability of receiving a diagnosis of albinism (*i.e.* the albinism risk) was estimated using data from the University Hospital of Bordeaux albinism cohort and the Genomics England 100,000 Genomes Project dataset. A case-control analysis of a binary trait (presence/absence of albinism) was conducted assuming a recessive model. Logistic regression using the Firth bias reduction method^37,38^ was utilised (as implemented in “logistf” R package). The following parameters were set:

- inclusion criteria: the analysis was conducted only on study subjects who: (i) were projected to have European-like ancestries, (ii) were not found to have genotypes in keeping with a molecular diagnosis of albinism, (iii) carried no pathogenic or likely pathogenic variants in the *TYR* or *OCA2* gene (with the exception of the two studied changes *TYR*:c.1205G>A and *OCA2*:c.1327G>A).
- covariates: sex; number of common albinism-associated alleles (see relevant sub-section below).

### Ancestry

To increase the genetic similarity between the studied case and control cohorts, we focused on individuals who were likely to fall within the same broad ancestral group. It is highlighted though that the utilised approach was imperfect as it was not possible to objectively determine genetic ancestry in individuals from the University Hospital of Bordeaux albinism cohort (as mentioned above, self-identified ethnicity was instead used as a surrogate). Confounding by ancestry (*i.e.* population stratification) is therefore a possibility.^39^ We attempted to quantify this by calculating the genomic inflation factor lambda (λGC). A total of 35 single-nucleotide variants were selected using a previously described approach (see Supplementary Table 7 and ^18^). Case-control comparisons were then made for each of these 35 λGC markers using Firth regression analysis. The resulting test statistics were used to calculate the median value of λGC.^18^

### Albinism-related genotypes

To reduce the likelihood of obtaining spurious results due to the impact of albinism-related variants other than the two studied changes (*TYR*:c.1205G>A and *OCA2*:c.1327G>A), we excluded individuals who carried genotypes consistent with a molecular diagnosis of albinism. This included study subjects with:

- homozygous pathogenic or likely pathogenic HGMD-listed variants in autosomal albinism-related genes *(TYR, OCA2, TYRP1, SLC45A2, SLC24A5, C10ORF11, HPS 1* to *10, LYST, SLC38A8*),
- presumed biallelic pathogenic or likely pathogenic HGMD-listed variants in autosomal albinism-related genes *(TYR, OCA2, TYRP1, SLC45A2, SLC24A5, C10ORF11, HPS 1* to *10, LYST, SLC38A8*),
- hemizygous pathogenic or likely pathogenic HGMD-listed variants in *GPR143* or *FRMD7,*
- the *TYR* c.[-301C;575C>A;1205G>A] or *TYR* c.[-301C;575C;1205G>A] albinism-related haplotype^19^ in homozygous state
- the *TYR* c.[-301C;575C>A;1205G>A] or *TYR* c.[-301C;575C;1205G>A] albinism-related haplotype in heterozygous state plus a heterozygous pathogenic or likely pathogenic variant in *TYR*.

We also excluded from our analysis individuals who were found to carry pathogenic or likely pathogenic variants in the *TYR* or *OCA2* gene (in heterozygous state). Small scale genetic changes (with the exception of *TYR*:c.1205G>A and *OCA2*:c.1327G>A), copy number and structural variants were factored in.

### Common albinism-associated alleles

The effect of one of the studied changes (*TYR* c.1205G>A) has been previously shown to be modified by the following two variants^19–21,25,34–36^ which have been added as a covariate to the regression model:

- *TYR* c.-301C>T [rs4547091], located in the *TYR* promoter; the derived, non-ancestral allele c.-301C (or c.-301=) has a MAF of ∼60% in non-Finnish European (NFE) populations in the Genome Aggregation Database (gnomAD v2.1.1)^33^;
- *TYR* c.575C>A (p.Ser192Tyr) [rs1042602], which has a MAF of ∼36% in gnomAD NFE populations.

It is highlighted that the *TYR* c.-301= and the *TYR* c.575C>A (p.Ser192Tyr) alleles are frequently found in linkage disequilibrium so the presence of the latter would, in most cases, suggest the presence of the former on the same haplotype.

### Case-control comparisons between TYR/OCA2 genotype groups

Our primary analysis focused on performing case-control comparisons between groups of genotypes that include different alleles at positions *TYR*:c.1205 and *OCA2*:c.1327. These groups were:

- group A (reference group): homozygous for *TYR*:c.1205= and *OCA2*:c.1327=
- group B (single TYR heterozygote group): heterozygous for *TYR*:c.1205G>A and homozygous for *OCA2*:c.1327=.
- group C (single OCA2 heterozygote group): homozygous for *TYR*:c.1205G= and heterozygous for *OCA2*:c.1327G>A.
- group D (dual heterozygote group): heterozygous for the *TYR*:c.1205G>A and *OCA2*:c.1327G>A variant combination.

Analyses were also conducted in individuals who were found to be homozygous for *TYR*:c.1205G>A (but are considered unsolved, *i.e.* they do not carry the *TYR* c.575C>A or the *TYR* c.-301= variant in the homozygous state); these were split into two groups:

- *group E:* homozygous for *TYR*:c.1205G>A and *OCA2*:c.1327=;
- *group F:* homozygous for *TYR*:c.1205G>A and heterozygous for OCA2:c.1327G>A

Group A was used as the reference group to which all other groups were compared using Firth regression analysis (*i.e.* the odds ratio for this group was fixed at 1).

The primary analysis focused on a mixed case cohort (including 174 probands from the University Hospital of Bordeaux cohort and 31 cases from the Genomics England 100,000 Genomes Project dataset) and a control cohort from the Genomics England 100,000 Genomes Project dataset (20,350 unrelated individuals) (Fig.2; Supplementary Fig. 1a; Supplementary Table 2). A secondary analysis focusing only on individuals from the Genomics England 100,000 Genomes Project dataset (31 cases, 20,350 controls) was also undertaken (Supplementary Fig. 1b; Supplementary Table 8).

### Replication of findings in the UK Biobank cohort

The effect of dual heterozygosity for *TYR:*c.1205G>A and *OCA2:*c.1327G>A was studied in the UK Biobank. UK Biobank is a biomedical resource containing in-depth genetic and health information from >500,000 individuals from across the UK.^40^ A subset of UK Biobank volunteers underwent enhanced phenotyping including visual acuity testing (131,985 individuals) and imaging of the central retina (84,748 individuals)^55^; the latter was obtained using optical coherence tomography (OCT), a non-invasive imaging test that rapidly generates cross-sectional retinal scans at micrometre-resolution.^56,57^ All UK Biobank volunteers whose data were analysed as part of this study were imaged using the 3D OCT-1000 Mark II device (Topcon, Japan); the relevant methodology has been previously described.^55^ Notably, the diagnosis of albinism has been assigned to 39 UK Biobank participants (ICD-10 term “Albinism” [E70.3] in data field 41270 or resource 591) (Supplementary Table 10).

Aiming to assess the robustness of the results in the combined University Hospital of Bordeaux and Genomics England 100,000 Genomes Project dataset, we performed case-control analyses in UK Biobank volunteers. We focused on individuals who were estimated by principal component analysis to have European-like ancestries (data field 22006). First, genotyping array data were used to obtain genotypes for the *TYR:*c.1205G>A variant and *OCA2:*c.1327G>A variant (data field 22418 including information from the Applied Biosystems UK Biobank Axiom Array containing 825,927 markers). Only individuals reliably genotyped at both sites were retained. Aiming to reduce the likelihood of obtaining spurious signals due to the presence of albinism-related variants other than the two studied changes (*TYR*:c.1205G>A and *OCA2*:c.1327G>A), we excluded UK Biobank participants whose genotyping array data (data field 22418) suggested that they carried an HGMD-listed variant in an albinism-related gene (Supplementary Data 4).

We used Firth logistic regression to perform case-control comparisons between genotype groups A-D. Sex and the number of common albinism-associated alleles were used as covariates (Fig.3a; Supplementary Fig. 2; Supplementary Table 3).

Given that reduced visual acuity and increased central retinal thickness (due to underdevelopment of the fovea) are two hallmark features of albinism we investigated the impact of selected *TYR/OCA2* variant combinations on these two quantitative traits. Aiming to reduce the likelihood of obtaining spurious signals due to the presence/impact of ophthalmic conditions/features not related to albinism, we excluded all UK Biobank volunteers that did not have the ICD-10 term “Albinism” [E70.3] and were assigned an ophthalmology-related ICD-10 code (“Chapter VII Diseases of the eye and adnexa”).

We first obtained data on the left LogMAR visual acuity for UK Biobank volunteers (data field 5201, “instance 0” datasets). These visual acuity measurements were then used to compare visual performance between groups of volunteers with selected *TYR/OCA2* genotype combinations (genotype groups A-D) (Supplementary Fig. 3). As the obtained distributions deviated from normality, the Kruskal-Wallis test was used. Pair-wise comparisons were performed and the p-values were adjusted using the Benjamini-Hochberg method (Fig.3b; Supplementary Table 4). Additional analyses using a linear regression approach were also conducted; sex and the number of common albinism-associated alleles were included as covariates (Supplementary Table 5).

We then obtained central retinal thickness measurements from UK Biobank OCT images using the “macular thickness at the central subfield” data field (27802; defined as the average distance between the hyperreflective bands corresponding to the RPE and the internal limiting membrane (ILM), across the central 1 mm diameter circle of the ETDRS grid) for UK Biobank volunteers.^58^ The obtained measurements were subsequently used to compare central retinal thickness between groups of UK Biobank volunteers with selected *TYR/OCA2* genotype combinations (groups A-D). As some of the obtained distributions deviated from normality, the Kruskal-Wallis test was used. Pair-wise comparisons were performed and the p-values were adjusted using the Benjamini-Hochberg method (Fig.3c; Supplementary Table 4). Additional analyses using a linear regression approach were also conducted; sex and the number of common albinism-associated alleles were included as covariates (Supplementary Table 6).

It is noted that we opted to perform all UK Biobank analyses on left eye data. This is because, as per study protocol^55^, the left eye was tested after the right eye, potentially making the measurements from the left side less prone to artifacts (as the participants were more familiar with the test).

### Additional analysis in the University Hospital of Bordeaux albinism cohort

Further analyses were undertaken using only data from individuals in the University Hospital of Bordeaux albinism cohort. After excluding study subjects who did not report having European-like ancestries, the cohort was split into cases with and cases without a molecular diagnosis of albinism (*i.e.* “solved” and “unsolved”). Individuals without a molecular diagnosis who carried heterozygous pathogenic or likely pathogenic variants in the *TYR* or *OCA2* gene (with the exception of the two studied changes *TYR*:c.1205G>A and *OCA2*:c.1327G>A) were then removed from the unsolved group. The patterns observed in genotype groups A-D were then assessed using both contingency table methods and Firth regression analyses (Supplementary Table 9).

### Ethics approval

Informed consent was obtained from all participants or their parents in the case of minors. The study of individuals from the University Hospital of Bordeaux albinism cohort has been approved by the relevant local ethics committee (Comité de Protection des Personnes Sud-Ouest et Outre Mer III, Bordeaux, France). The informed consent process for the Genomics England 100,000 Genomes Project has been approved by the National Research Ethics Service Research Ethics Committee for East of England – Cambridge South Research Ethics Committee. The UK Biobank has received approval from the National Information Governance Board for Health and Social Care and the National Health Service North West Centre for Research Ethics Committee (Ref: 11/NW/0382). All investigations were conducted in accordance with the tenets of the Declaration of Helsinki.

## DATA AVAILABILITY

The Genomics England 100,000 Genomes Project dataset is available under restricted access through a procedure described at https://www.genomicsengland.co.uk/about-gecip/for-gecip-members/data-and-data-access. The UK Biobank dataset is available under restricted access through a procedure described at http://www.ukbiobank.ac.uk/using-the-resource/. All other data supporting the findings of this study are available within the article (including its Supplementary Information files).

## CODE AVAILABILITY

The scripts used to analyse the datasets included in this study are available at https://github.com/davidjohngreen/tyr-oca2.

## ACKNOWLEDGEMENTS

We acknowledge the following sources of funding: the Wellcome Trust (224643/Z/21/Z, Clinical Research Career Development Fellowship to P.I.S.; 200990/Z/16/Z, Transforming Genetic Medicine Initiative to G.C.B.); the UK National Institute for Health Research (NIHR) Clinical Lecturer Programme (CL-2017-06-001 to P.I.S.); Retina UK and Fight for Sight (GR586, RP Genome Project - UK Inherited Retinal Disease Consortium to G.C.B.); Christopher Green (D.J.G.); the French Albinism Association (Genespoir) and the French National Research Agency (Agence Nationale de la Recherche; ANR-21-CE17-0041-01 to B.A.).

The UK Biobank Eye and Vision Consortium is supported by funding from the NIHR Biomedical Research Centre at Moorfields Eye Hospital and UCL Institute of Ophthalmology, the Alcon Foundation and the Desmond Foundation. The complete list of members of this Consortium can be found in the Supplementary Information This research was made possible through access to the data and findings generated by the 100,000 Genomes Project. The 100,000 Genomes Project is managed by Genomics England Limited (a wholly owned company of the Department of Health and Social Care). The 100,000 Genomes Project is funded by the NIHR and NHS England. The Wellcome Trust, Cancer Research UK and the Medical Research Council have also funded research infrastructure. The 100,000 Genomes Project uses data provided by patients and collected by the National Health Service (NHS) as part of their care and support. We acknowledge the contribution of the Genomics England Research Consortium to the 100,000 Genomes Project. The complete list of members of this Consortium can be found in the Supplementary Information.

Lastly, we acknowledge the help of Cécile Courdier at the University Hospital of Bordeaux Molecular Genetics Laboratory, Sophie Javerzat at the University of Bordeaux, Dave Gerrard at the University of Manchester, and Claire Hardcastle, Chris Campbell, Steph Barton and Simon Ramsden at the North West of England Genomic Laboratory Hub.

## AUTHOR CONTRIBUTIONS STATEMENT

P.I.S. conceived and designed the experiments. The UK Biobank Eye and Vision Consortium, T.F., E.B., B.A., G.C.B. and P.I.S provided datasets and analytical tools. D.J.G., V.M., E.L., C.P., T.F., B.A. and P.I.S. analysed the data. P.I.S. wrote the manuscript with support from D.J.G. All authors critically revised and approved the manuscript.

G.C.B. and B.A. contributed equally to this work.

## COMPETING INTERESTS STATEMENT

E.B. is a paid consultant and equity holder of Oxford Nanopore, a paid consultant to Dovetail, and a non-executive director of Genomics England, a limited company wholly owned by the UK Department of Health and Social Care. All other authors declare no competing interests.

## UK Biobank Eye and Vision Consortium

Graeme C. Black^1,5^, Panagiotis I. Sergouniotis^1,4,5,6^

1 Division of Evolution, Infection and Genomics, School of Biological Sciences, Faculty of Biology, Medicine and Health, University of Manchester, Manchester, UK.

4 European Molecular Biology Laboratory, European Bioinformatics Institute (EMBL-EBI), Wellcome Genome Campus, Cambridge, UK.

5 Manchester Centre for Genomic Medicine, Saint Mary’s Hospital, Manchester University NHS Foundation Trust, Manchester, UK.

6 Manchester Royal Eye Hospital, Manchester University NHS Foundation Trust, Manchester, UK.

A full list of members appears in the Supplementary Information.

## Notes

### Author Declarations

Informed consent was obtained from all participants or their parents in the case of minors. The study of individuals from the University Hospital of Bordeaux albinism cohort has been approved by the relevant local ethics committee (Comite de Protection des Personnes Sud-Ouest et Outre Mer III, Bordeaux, France). The informed consent process for the Genomics England 100,000 Genomes Project has been approved by the National Research Ethics Service Research Ethics Committee for East of England - Cambridge South Research Ethics Committee. The UK Biobank has received approval from the National Information Governance Board for Health and Social Care and the National Health Service North West Centre for Research Ethics Committee (Ref: 11/NW/0382). All investigations were conducted in accordance with the tenets of the Declaration of Helsinki.

### Summary of Updates

In this revised version we have included comprehensive UK Biobank re-analyses that strengthen the finding of the replication study. In addition, minor changes were made to the figures and tables.

## REFERENCES

1. Boycott, K.M., et al. A diagnosis for all rare genetic diseases: The horizon and the next frontiers. Cell.177, 32–37 (2019). doi: 10.1016/j.cell.2019.02.040.

2. Splinter, K., et al. Effect of genetic diagnosis on patients with previously undiagnosed disease. N Engl J Med. 379, 2131–2139 (2018). doi: 10.1056/NEJMoa1714458.

3. Posey, J.E., et al. Insights into genetics, human biology and disease gleaned from family based genomic studies. Genet Med. 21, 798–812 (2019). doi: 10.1038/s41436-018-0408-7.

4. Zurek, B., et al. Solve-RD: systematic pan-European data sharing and collaborative analysis to solve rare diseases. Eur J Hum Genet. 29, 1325–1331 (2021). doi: 10.1038/s41431-021-00859-0

5. 100,000 Genomes Project Pilot Investigators. 100,000 Genomes Pilot on rare-disease diagnosis in health care - Preliminary report. N Engl J Med. 385, 1868-1880 (2021). doi: 10.1056/NEJMoa2035790.

6. Boycott, K.M., et al. Care4Rare Canada: Outcomes from a decade of network science for rare disease gene discovery. Am J Hum Genet. 109, 1947–1959 (2022). doi: 10.1016/j.ajhg.2022.10.002.

7. Schäffer, A.A. Digenic inheritance in medical genetics. J Med Genet. 50, 641–652 (2013). doi: 10.1136/jmedgenet-2013-101713.

8. Katsanis, N. The continuum of causality in human genetic disorders. Genome Biol. 17, 233 (2016). doi: 10.1186/s13059-016-1107-9.

9. Deltas C. Digenic inheritance and genetic modifiers. Clin Genet. 93, 429–438 (2018). doi: 10.1111/cge.13150.

10. Nachtegael, C., et al. Scaling up oligogenic diseases research with OLIDA: the Oligogenic Diseases Database. Database (Oxford). 2022, baac023 (2022). doi: 10.1093/database/baac023.

11. Burkard, M. et al. Accessory heterozygous mutations in cone photoreceptor CNGA3 exacerbate CNG channel-associated retinopathy. J Clin Invest. 128, 5663–5675 (2018). doi: 10.1172/JCI96098

12. Cooper, D.N., Krawczak, M., Polychronakos, C., Tyler-Smith, C. & Kehrer-Sawatzki, H. Where genotype is not predictive of phenotype: towards an understanding of the molecular basis of reduced penetrance in human inherited disease. Hum Genet. 132, 1077–1130 (2013). doi: 10.1007/s00439-013-1331-2.

13. Kousi, M. & Katsanis, N. Genetic modifiers and oligogenic inheritance. Cold Spring Harb Perspect Med. 5, a017145 (2015). doi: 10.1101/cshperspect.a017145.

14. Costanzo, M., et al. Global genetic networks and the genotype-to-phenotype relationship. Cell. 177, 85–100 (2019). doi: 10.1016/j.cell.2019.01.033.

15. Mukherjee, S., et al. Identifying digenic disease genes via machine learning in the Undiagnosed Diseases Network. Am J Hum Genet. 108, 1946–1963 (2021). doi: 10.1016/j.ajhg.2021.08.010.

16. Papadimitriou, S., et al. Predicting disease-causing variant combinations. Proc Natl Acad Sci U S A. 116:11878–11887 (2019). doi: 10.1073/pnas.1815601116.

17. Brehm, A., et al. Additive loss-of-function proteasome subunit mutations in CANDLE/PRAAS patients promote type I IFN production. J Clin Invest. 125, 4196–4211 (2015). doi: 10.1172/JCI81260.

18. Bakker, R., et al. The retinal pigmentation pathway in human albinism: Not so black and white. Prog Retin Eye Res. 91, 101091 (2022). doi: 10.1016/j.preteyeres.2022.101091.

19. Michaud, V., et al. The contribution of common regulatory and protein-coding TYR variants to the genetic architecture of albinism. Nat Commun. 13, 3939 (2022). doi: 10.1038/s41467-022-31392-3.

20. Campbell, P., et al. Clinical and genetic variability in children with partial albinism. Sci Rep. 9, 16576 (2019). doi: 10.1038/s41598-019-51768-8.

21. Monfermé, S., et al. Mild form of oculocutaneous albinism type 1: phenotypic analysis of compound heterozygous patients with the R402Q variant of the *TYR* gene. Br J Ophthalmol. 103, 1239–1247 (2019). doi: 10.1136/bjophthalmol-2018-312729

22. Lasseaux, E., et al. Molecular characterization of a series of 990 index patients with albinism. Pigment Cell Melanoma Res. 31, 466–474 (2018). doi: 10.1111/pcmr.12688.

23. Caulfield, M., et al. The National Genomic Research Library. 10.6084/m9.figshare.4530893.v7 (2017).

24. Okazaki, A. & Ott, J. Machine learning approaches to explore digenic inheritance. Trends Genet. 38, 1013–1018 (2022). doi: 10.1016/j.tig.2022.04.009.

25. Jagirdar, K., et al. Molecular analysis of common polymorphisms within the human tyrosinase locus and genetic association with pigmentation traits. Pigment Cell Melanoma Res. 27, 552–564 (2014). doi: 10.1111/pcmr.12253.

26. Dolinska, M.B., et al. Oculocutaneous albinism type 1: link between mutations, tyrosinase conformational stability, and enzymatic activity. Pigment Cell Melanoma Res. 30, 41–52 (2017). doi: 10.1111/pcmr.12546.

27. Sviderskaya, E.V., et al. Complementation of hypopigmentation in p-mutant (pink-eyed dilution) mouse melanocytes by normal human P cDNA, and defective complementation by OCA2 mutant sequences. J Invest Dermatol. 108, 30–34 (1997). doi: 10.1111/1523-1747.ep12285621

28. Bellono, N.W., Escobar, I.E., Lefkovith, A.J., Marks, M.S. & Oancea, E. An intracellular anion channel critical for pigmentation. Elife. 3, e04543 (2014). doi: 10.7554/eLife.04543.

29. Bellono, N.W., Escobar, I.E. & Oancea, E. A melanosomal two-pore sodium channel regulates pigmentation. Sci Rep. 6, 26570 (2016). doi: 10.1038/srep26570.

30. Pavan W.J. & Sturm, R.A. The genetics of human skin and hair pigmentation. Annu Rev Genomics Hum Genet. 20, 41–72 (2019). doi: 10.1146/annurev-genom-083118-015230.

31. Landrum, M.J., et al. ClinVar: improving access to variant interpretations and supporting evidence. Nucleic Acids Res. 46, D1062–D1067 (2018). doi: 10.1093/nar/gkx1153. [accessions VCV000003779.25 & VCV000000955.52].

32. Buniello, A., et al. The NHGRI-EBI GWAS Catalog of published genome-wide association studies, targeted arrays and summary statistics 2019. Nucleic Acids Res. 47, D1005–D1012 (2019). doi: 10.1093/nar/gky1120. [entries for rs1126809 and rs74653330]

33. Karczewski, K.J., et al. The mutational constraint spectrum quantified from variation in 141,456 humans. Nature. 581, 434–443 (2020). doi: 10.1038/s41586-020-2308-7. [entries for 11-89017961-G-A (GRCh37) and 15-28228553-C-T (GRCh37)]

34. Loftus, S.K., et al. Haplotype-based analysis resolves missing heritability in oculocutaneous albinism type 1B. Am J Hum Genet. S0002-9297, 00169-6 (2023). doi: 10.1016/j.ajhg.2023.05.012.

35. Lin, S., et al. Evidence that the Ser192Tyr/Arg402Gln in *cis* Tyrosinase gene haplotype is a disease-causing allele in oculocutaneous albinism type 1B (OCA1B). NPJ Genom Med. 7, 2 (2022). doi: 10.1038/s41525-021-00275-9.

36. Grønskov, K., et al. A pathogenic haplotype, common in Europeans, causes autosomal recessive albinism and uncovers missing heritability in OCA1. Sci Rep. 9, 645 (2019). doi: 10.1038/s41598-018-37272-5.

37. Heinze, G. & Schemper, M. A solution to the problem of separation in logistic regression. Stat Med. 21, 2409–19 (2002). doi: 10.1002/sim.1047.

38. Firth, D. Bias reduction of maximum likelihood estimates. Biometrika. 80, 27–38 (1993). doi: 10.1093/biomet/80.1.27

39. Marchini, J., Cardon, L.R., Phillips, M.S. & Donnelly, P. The effects of human population structure on large genetic association studies. Nat Genet. 36, 512–517 (2004). doi: 10.1038/ng1337

40. Bycroft, C., et al. The UK Biobank resource with deep phenotyping and genomic data. Nature. 562, 203–209 (2018). doi: 10.1038/s41586-018-0579-z.

41. Masson, E., et al. Expanding ACMG variant classification guidelines into a general framework. Hum Genomics. 16, 31 (2022). doi: 10.1186/s40246-022-00407-x.

42. Richards, S., et al. Standards and guidelines for the interpretation of sequence variants: a joint consensus recommendation of the American College of Medical Genetics and Genomics and the Association for Molecular Pathology. Genet Med. 17, 405–424 (2015). doi: 10.1038/gim.2015.30.

43. Freedman, M.L., et al. Assessing the impact of population stratification on genetic association studies. Nat Genet. 36, 388–93 (2004). doi: 10.1038/ng1333.

44. Devlin, B. & Roeder, K. Genomic control for association studies. Biometrics. 55, 997–1004 (1999). doi: 10.1111/j.0006-341x.1999.00997.x

45. Dadd, T., Weale, M.E. & Lewis, C.M. A critical evaluation of genomic control methods for genetic association studies. Genet Epidemiol. 33, 290–298 (2009). doi: 10.1002/gepi.20379.

46. Turnbull, C. et al. The 100,000 Genomes Project: bringing whole genome sequencing to the NHS. BMJ 361, 1687 (2018). doi: 10.1136/bmj.k1687

47. Köhler, S. et al. The Human Phenotype Ontology in 2021. Nucleic Acids Res. 49, D1207– D1217 (2021). doi: 10.1093/nar/gkaa1043

48. Raczy, C. et al. Isaac: ultra-fast whole-genome secondary analysis on Illumina sequencing platforms. Bioinformatics 29, 2041–2043 (2013). doi: 10.1093/bioinformatics/btt314.

49. Chen, X., et al. Manta: rapid detection of structural variants and indels for germline and cancer sequencing applications. Bioinformatics 32, 1220–1222 (2016). doi: 10.1093/bioinformatics/btv710.

50. Roller, E., Ivakhno, S., Lee, S., Royce, T. & Tanner, S. Canvas: versatile and scalable detection of copy number variants. Bioinformatics 32, 2375–2377 (2016). doi: 10.1093/bioinformatics/btw163.

51. Tan, A., Abecasis, G.R. & Kang, H.M. Unified representation of genetic variants. Bioinformatics 31, 2202–2204 (2015). doi: 10.1093/bioinformatics/btv112

52. Danecek, P. et al. Twelve years of SAMtools and BCFtools. Gigascience 10, giab008 (2021).

53. McLaren, W. et al. The Ensembl Variant Effect Predictor. Genome Biol. 17, (2016). doi: 10.1186/s13059-016-0974-4

54. Stenson, P.D., et al. The Human Gene Mutation Database (HGMD^®^): optimizing its use in a clinical diagnostic or research setting. Hum Genet. 139, 1197–1207 (2020). doi: 10.1007/s00439-020-02199-3.

55. Chua, S.Y.L., et al. Cohort profile: design and methods in the eye and vision consortium of UK Biobank. BMJ Open 9, e025077 (2019).

56. Bouma, B.E., et al. Optical coherence tomography. Nat Rev Methods Primers. 2, 79 (2022). doi: 10.1038/s43586-022-00162-2.

57. Drexler, W. & Fujimoto, J.G. State-of-the-art retinal optical coherence tomography. Prog Retin *Eye* *Res*. 27, 45–88 (2008). doi: 10.1016/j.preteyeres.2007.07.005

58. Currant, H., et al. Genetic variation affects morphological retinal phenotypes extracted from UK Biobank optical coherence tomography images. PLoS Genet. 17, e1009497 (2021). doi: 10.1371/journal.pgen.1009497

