## Supplementary Information for "The co-occurrence of genetic variants in the *TYR* and *OCA2* genes confers susceptibility to albinism"

### SUPPLEMENTARY FIGURES

**Supplementary Figure 1**. Analyses in selected subsets of the studied cohorts suggest that dual heterozygosity for *TYR*:c.1205G>A (p.Arg402Gln) and *OCA2*:c.1327G>A (p.Val443Ile) is associated with an increased probability of receiving an albinism diagnosis.

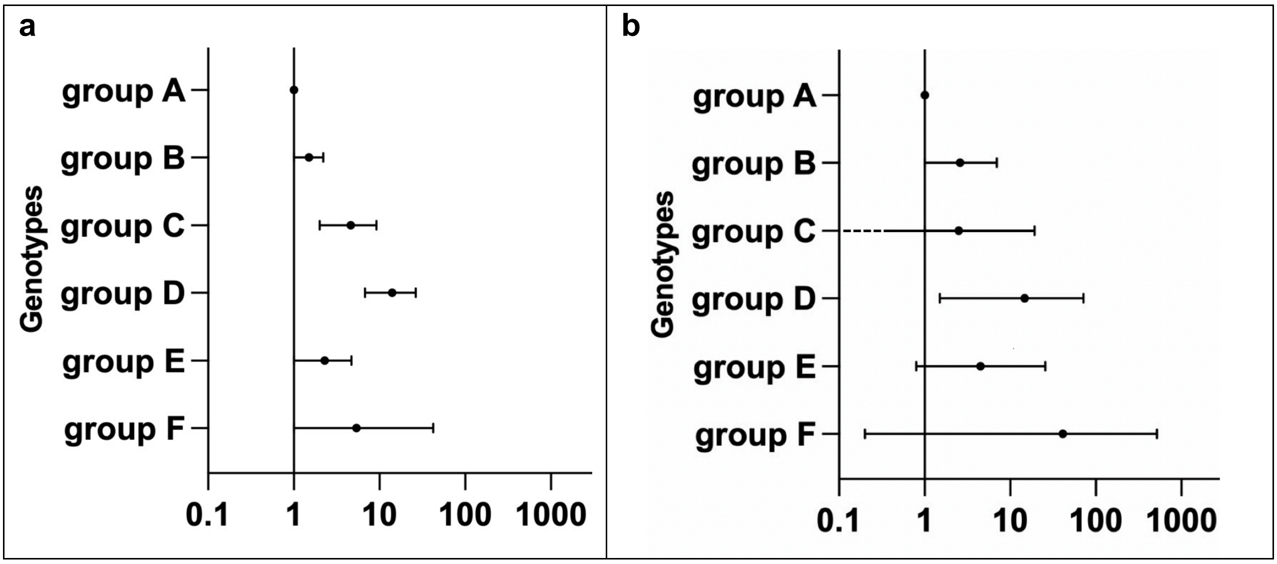


The x-axes are showing odds ratios corresponding to the probability of receiving a diagnosis of albinism. The circle in the middle of each horizontal line (95% confidence interval) represents the point estimate of each odds ratio. A $\log_{10}$ scale with 1 as the reference point is used.

Both panels outline the findings of case-control analyses using Firth regression. These analyses were conducted only on study subjects who: (i) were projected to have European-like ancestries, (ii) were not found to have genotypes in keeping with a molecular diagnosis of albinism, (iii) carried no pathogenic or likely pathogenic variants in the *TYR* or *OCA2* gene (with the exception of the two studied changes *TYR*:c.1205G>A and *OCA2*:c.1327G>A).

The following genotype groups were studied:

- ***group A*** (reference group): homozygous for *TYR*:c.1205= and *OCA2*:c.1327=.
- ***group B*** (single *TYR* heterozygote group): heterozygous for *TYR*:c.1205G>A and homozygous for *OCA2*:c.1327=.
- ***group C*** (single *OCA2* heterozygote group): homozygous for *TYR*:c.1205= and heterozygous for *OCA2*:c.1327G>A.
- ***group D*** (dual heterozygote group): heterozygous for *TYR*:c.1205G>A and *OCA2*:c.1327G>A.

Findings in individuals who were homozygous for *TYR*:c.1205G>A (but are considered unsolved, *i.e.* they do not carry the *TYR* c.575C>A or the *TYR* c.-301= variant in the homozygous state) are also shown; these were split into two groups:

- ***group E:*** homozygous for *TYR*:c.1205G>A and *OCA2*:c.1327=;
- ***group F:*** homozygous for *TYR*:c.1205G>A and heterozygous for OCA2:c.1327G>A

Panel **a**, shows the results obtained following the primary analysis which focused on a mixed case cohort (including 173 probands from the University Hospital of Bordeaux cohort and 31 cases from the 100K_GP dataset) and a control cohort from the 100K_GP dataset (20,350 unrelated individuals). This graph is an extension of that shown in Fig.2 and is included here to facilitate comparison with the graph in panel b. Further information and relevant numerical data can be found in Supplementary Table 2.

Panel **b**, shows the results obtained following a secondary analysis focusing on cases and controls from the 100K_GP dataset only. Further information and relevant numerical data can be found in Supplementary Table 8.

Overall, the findings of the secondary analysis (which includes case and control populations that can be considered genetically well-mixed) are well aligned with those of the primary study.

100K_GP corresponds to Genomics England 100,000 Genomes Project.

**Supplementary Figure 2**. Flowchart outlining the UK Biobank case-control study and visual acuity and retinal thickness analyses

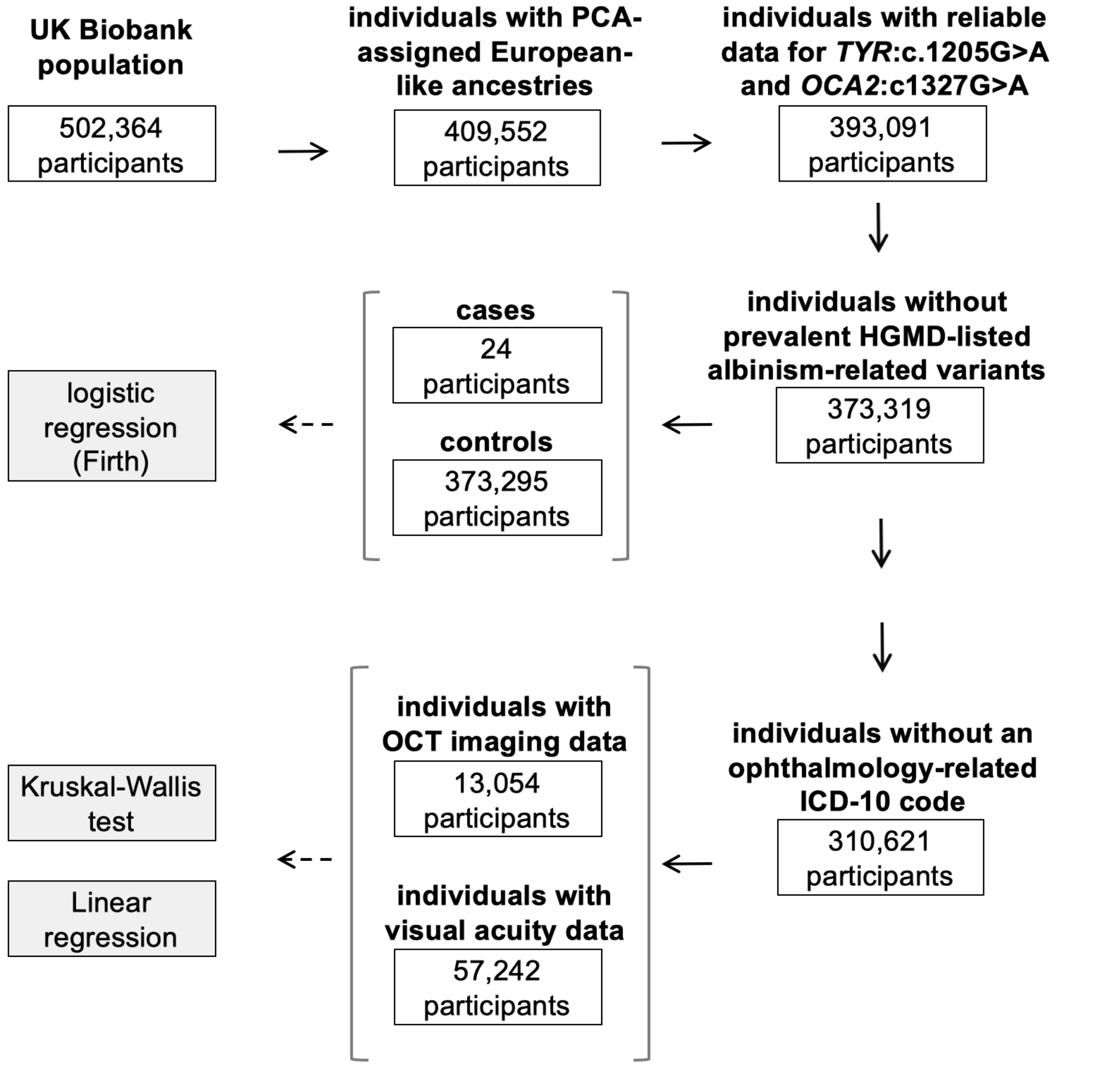


Aiming to validate the findings of the primary analysis (Fig.1, Supplementary Fig.1a, Supplementary Table 2), we performed additional studies in an independent cohort, the UK Biobank.

To reduce the likelihood of obtaining spurious signals due to population stratification effects or due to the presence of albinism-related variants other than the two studied changes (*TYR*:c.1205G>A and *OCA2*:c.1327G>A), we focused only on individuals who were projected to have European-like ancestries and whose genotyping array data suggested that they did not carry an HGMD-listed variant in an albinism-related gene. The following two covariates were used in logistic and linear regression analyses: sex and number of common albinism-associated alleles, *i.e.* DNA sequence alterations in the genomic locations corresponding to *TYR*:c.−301C>T [rs4547091] and *TYR*:c.575C>A (p.Ser192Tyr) [rs1042602]. These two variants have been previously shown to modify the effect of *TYR*:c.1205G>A which is a common missense change that can act as a “hypomorphic” variant.

The relevant results can be found in Fig.3a-c and in Supplementary Tables 4-6.

PCA, principal component analysis; OCT, optical coherence tomography; HGMD-listed variants, variants with a “total” minor allele frequency (MAF) <1% in the Genome Aggregation Database (gnomAD v2.1.1) and a “disease-causing” (DM) label in the Human Gene Mutation Database (HGMD) v2021.2.

**Supplementary Figure 3**. Segregation analysis data in four illustrative families that include individuals with albinism and dual heterozygosity for *TYR*:c.1205G>A (p.Arg402Gln) and *OCA2*:c.1327G>A (p.Val443Ile).

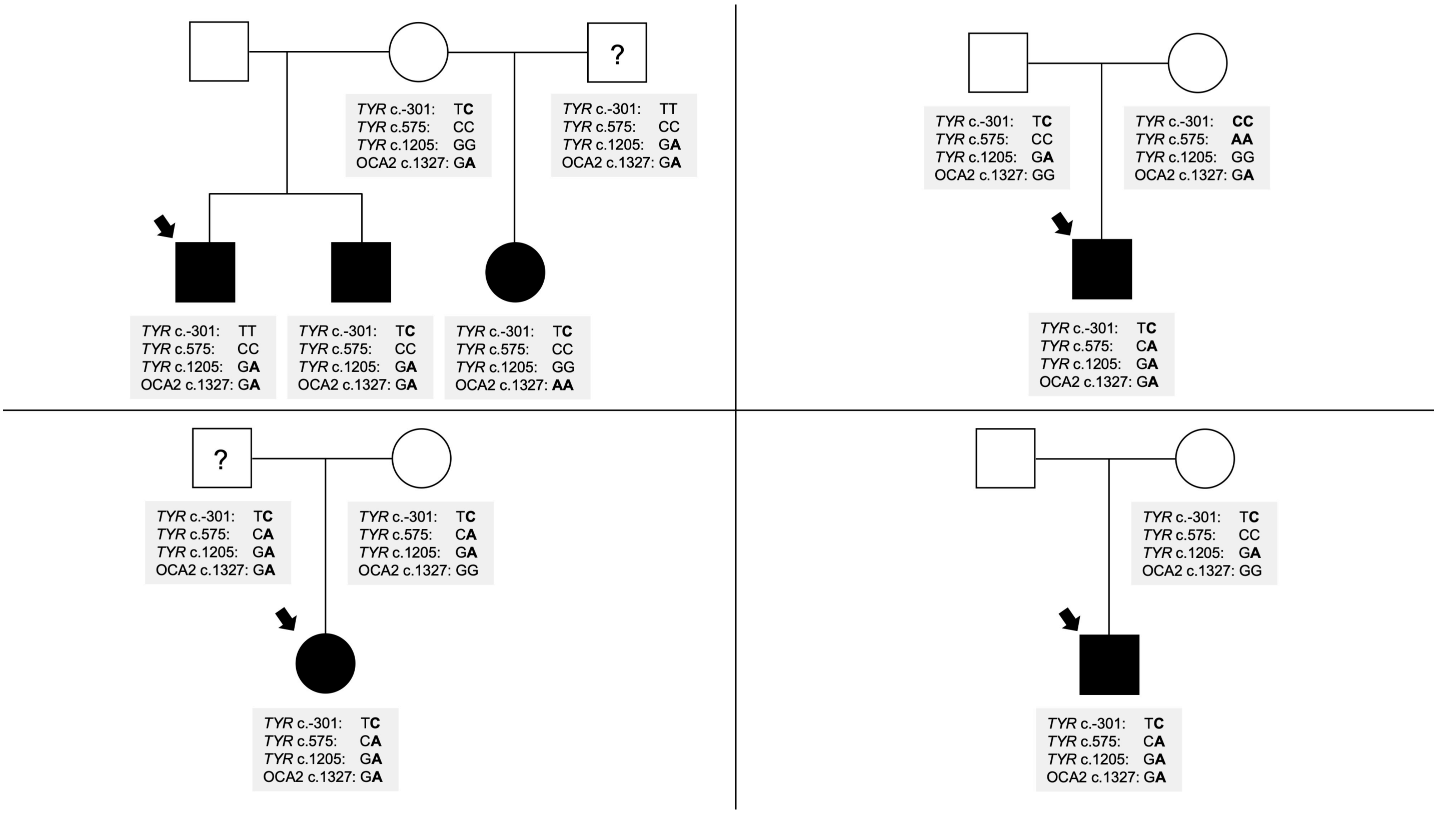


Arrows are used to indicate the proband in each family. The question mark (?) symbol indicates unknown affection status. It is highlighted that dual heterozygosity for *TYR*:c.1205G>A (p.Arg402Gln) and *OCA2*:c.1327G>A (p.Val443Ile) should not be viewed as a genotype acting in a Mendelian pattern. Many individuals carrying this variant combination are going to be unaffected (or subclinically affected) and this reflects the fact that this genotype acts as a probabilistic marker of albinism. It can be speculated that the expressivity/penetrance of dual heterozygosity for *TYR*:c.1205G>A and *OCA2*:c.1327G>A can be modified by other non-genetic or genetic factors (such as the presence of *TYR*:c.575C>A (p.Ser192Tyr)); studies in large, comprehensively phenotyped cohorts are expected to provide further insights into more complex variant interaction patterns.

### SUPPLEMENTARY TABLES

**Supplementary Table 1**. Cohort characteristics

| | **cases** | **controls** |
|---|---|---|
| number of probands | 1,015 + 105 [a] | 29,451 |
| percentage of female probands | 44% [b] | 48% |
| percentage of probands with European-like ancestries [c] | 84% | 78% |
| genetic testing approach | gene panel testing or genome sequencing [a] | genome sequencing |

[a] 1,015 probands with a diagnosis of albinism were identified through the University Hospital of Bordeaux Molecular Genetics Laboratory database; most of these cases underwent gene panel testing. A further 105 affected probands were identified in the Genomics England 100,000 Genomes Project dataset; these cases underwent genome sequencing using a similar pipeline to the control cohort.

[b] The observed enrichment in male probands in the case cohort is due to the contribution of X-linked forms of albinism.

[c] Ancestry was estimated using a questionnaire in most cases (1,015/1,120) and using principal component analysis in the remaining cases and in controls.

**Supplementary Table 2**. Case-control comparisons between different groups of genotypes linked to the *TYR*:c.1205G>A (p.Arg402Gln) and *OCA2*:c.1327G>A (p.Val443Ile) variants:

University Hospital of Bordeaux Molecular Genetics Laboratory and Genomics England 100,000 Genome Project dataset analysis [a]

| **genotype** | | | | | | |
|---|---|---|---|---|---|---|
| ***TYR:* c.1205G>A (p.Arg402Gln) [rs1126809]** | ***OCA2*: c.1327G>A (p.Val443Ile) [rs74653330]** | **group** | **odds ratio** [b] | **95% confidence interval** | **unadjusted p-value** | **number of probands (cases / controls)** [a] |
| GG | GG | *A* | 1 | not applicable | | 103 / 10,369 |
| AG | GG | *B* | 1.5 | 1.0 – 2.2 | 0.044 | 71 / 8,011 |
| GG | AG | *C* | 4.6 | 2.0 – 9.2 | 0.002 | 7 / 158 |
| AG | AG | *D* | 12.8 | 6.0 – 24.7 | $2.1 \times 10^{-8}$ | 10 / 128 |
| AA [c] | GG | *E* | 2.2 | 1.0 – 4.6 | 0.04 | 13 / 1,658 |
| AA [c] | AG | *F* | 5.3 | 0 – 41.7 | 0.4 | 0 / 26 |
| GG | AA | | these genotypes were not studied as individuals in these groups are considered to have a molecular diagnosis (*i.e.* to be "solved") and therefore did not meet the inclusion criteria for analysis | | | |
| AG | AA | | | | | |
| AA | AA | | | | | |

[a] This analysis was conducted only on study subjects who: (i) were projected to have European-like ancestries, (ii) were not found to have genotypes in keeping with a molecular diagnosis of albinism, (iii) carried no pathogenic or likely pathogenic variants in the *TYR* or *OCA2* gene (with the exception of the two studied changes *TYR*:c.1205G>A and *OCA2*:c.1327G>A). Ancestry was estimated using a questionnaire in most cases (174/205) and using principal component analysis in the remaining cases and in controls.

[b] Firth logistic regression was used to generate odds ratios and unadjusted p-values; 204 probands with albinism and 20,350 unrelated controls were included in this analysis. Sex and the number of common albinism-associated alleles were used as covariates. The presented odds ratio corresponds to the probability of receiving a diagnosis of albinism. Group A was used as the reference group to which all other groups were compared to; the odds ratio for this group was therefore fixed at 1.

[c] As only individuals without a molecular diagnosis were considered, these groups include study subjects who were homozygous for *TYR*:c.1205G>A but did not carry the *TYR* c.-301= or the *TYR* c.575C>A variant in the homozygous state.

It is noted that the *TYR*:c.1205G>A and *OCA2*:c.1327G>A variants appear to have unequal effects. This is unsurprising given the fact that the common *TYR*:c.1205G>A change is considered to be a "hypomorphic" variant associated with relatively mild forms of albinism.

Although the above observations can be viewed as suggestive of a non-additive interaction between *TYR*:c.1205G>A and *OCA2*:c.1327G>A, such speculation would be inappropriate in the absence of focused analyses in studies with larger sample sizes.

A plot of these results can be found in Fig.2 and Supplementary Fig.1a.

**Supplementary Table 3**. Case-control comparisons between different groups of genotypes linked to the *TYR*:c.1205G>A (p.Arg402Gln) and *OCA2*:c.1327G>A (p.Val443Ile) variants:

UK Biobank dataset analysis [a]

| genotype | | | odds ratio [b] | 95% confidence interval | unadjusted p-value | number of probands (cases / controls) [a] |
|---|---|---|---|---|---|---|
| ***TYR:*** **c.1205G>A (p.Arg402Gln) [rs1126809]** | ***OCA2*****: c.1327G>A (p.Val443Ile) [rs74653330]** | **group** | | | | |
| GG | GG | *A* | 1 | not applicable | | 9 / 177,357 |
| AG | GG | *B* | 3.4 | 1.0 – 11.5 | 0.047 | 4 / 155,850 |
| GG | AG | *C* | 18.0 | 3.4 – 67.3 | 0.0026 | 2 / 3,322 |
| AG | AG | *D* | 42.8 | 6.9 – 196.5 | 0.0004 | 2 / 2,908 |

[a] In contrast to the primary analysis presented in Fig.2, Supplementary Fig.1a and Supplementary Table 2, this replication study focused on individuals (both cases and controls) from a different cohort, the UK Biobank. We only analyzed the subset of UK Biobank volunteers who were predicted by principal component analysis to have European-like ancestries and whose genotyping array data suggested that they did not carry an HGMD-listed variant in an albinism-related gene.

[b] Firth logistic regression was used to generate odds ratios and unadjusted p-values; 24 individuals with albinism and 373,295 controls were included in this analysis. Sex and the number of common albinism-associated alleles were used as covariates. The presented odds ratio corresponds to the probability of receiving a diagnosis of albinism. Group A was used as the reference group to which all other groups were compared; the odds ratio for this group was therefore fixed at 1.

It is noted that the UK Biobank cohort is not representative of the corresponding general population and the obtained absolute values and effect sizes should be interpreted with caution.

A plot of these results can be found in Fig.3a.

**Supplementary Table 4**. Visual acuity and central retinal thickness in different groups of genotypes linked to the *TYR*:c.1205G>A (p.Arg402Gln) and *OCA2*:c.1327G>A (p.Val443Ile) variants:

UK Biobank dataset analysis – nonparametric hypothesis testing

| genotype | | | Mean LogMAR visual acuity | Median LogMAR visual acuity | Mean central retinal thickness | Median central retinal thickness |
|---|---|---|---|---|---|---|
| ***TYR:*** **c.1205G>A (p.Arg402Gln) [rs1126809]** | ***OCA2*****: c.1327G>A (p.Val443Ile) [rs74653330]** | **group** | | | | |
| GG | GG | *A* | 0.0073 | -0.04 | 266 | 265 |
| GG | AG | *B* | 0.0069 | -0.04 | 265 | 264 |
| GG | AG | *C* | 0.0269 | -0.02 | 269 | 269 |
| AG | AG | *D* | 0.0314 | -0.02 | 271 | 271 |

The average ranks of groups A, B, C and D were compared using the Kruskal-Wallis method; a p-value of $1.4 \times 10^{-3}$ was obtained for visual acuity and a p-value of $8.2 \times 10^{-3}$ was obtained for central retinal thickness. Pair-wise comparisons between group D (*i.e.* dual heterozygosity for *TYR*:c.1205G>A and *OCA2*:c.1327G>A) and the other categories revealed the following Benjamini-Hochberg adjusted p-values:

VISUAL ACUITY:

- group D vs group A, p-value 0.0089
- group D vs group B, p-value 0.0089
- group D vs group C, p-value 0.67
- group D vs all other (*i.e.* non-group D) genotypes, p-value 0.0025

CENTRAL RETINAL THICKNESS:

- group D vs group A, p-value 0.045
- group D vs group B, p-value 0.045
- group D vs group C, p-value 0.47
- group D vs all other (*i.e.* non-group D) genotypes, p-value 0.0089

A plot of these results can be found in Fig.3b,c.

**Supplementary Table 5**. Visual acuity in different groups of genotypes linked to the *TYR*:c.1205G>A (p.Arg402Gln) and *OCA2*:c.1327G>A (p.Val443Ile) variants:

UK Biobank dataset analysis – linear regression

| **genotype** | | | | | | |
|---|---|---|---|---|---|---|
| ***TYR:* c.1205G>A (p.Arg402Gln) [rs1126809]** | ***OCA2*: c.1327G>A (p.Val443Ile) [rs74653330]** | **group** | **estimate** | **standard error** | **t-value** | **p-value (Benjamini-Hochberg adjusted)** |
| GG | GG | *A* | not applicable | | | |
| AG | GG | *B* | -0.0006 | 0.0021 | -0.29 | 0.775 |
| GG | AG | *C* | 0.0193 | 0.0093 | 2.07 | 0.057 |
| AG | AG | *D* | 0.0235 | 0.0097 | 2.42 | 0.015 |

The visual acuity between groups A, B, C and D was compared using linear regression with group A set as the reference. Sex and number of common albinism-associated alleles were used as covariates (Methods, Supplementary Fig.4). A p-value of $2.1 \times 10^{-15}$ was obtained for the model overall.

The number of individuals in the A, B, C, and D groups were as follows: 30,098; 26,190; 499; and 460.

**Supplementary Table 6**. Central retinal thickness in different groups of genotypes linked to the *TYR*:c.1205G>A (p.Arg402Gln) and *OCA2*:c.1327G>A (p.Val443Ile) variants:

UK Biobank dataset analysis [a] – linear regression

| **genotype** | | | | | | |
|---|---|---|---|---|---|---|
| ***TYR:* c.1205G>A (p.Arg402Gln) [rs1126809]** | ***OCA2*: c.1327G>A (p.Val443Ile) [rs74653330]** | **group** | **estimate** | **standard error** | **t-value** | **p-value (Benjamini-Hochberg adjusted)** |
| GG | GG | *A* | not applicable | | | |
| AG | GG | *B* | 0.4917 | 0.4653 | 1.06 | 0.290 |
| GG | AG | *C* | 2.1016 | 2.0475 | 1.03 | 0.305 |
| AG | AG | *D* | 7.1795 | 2.1953 | 3.27 | 0.001 |

The visual acuity between groups A, B, C and D was compared using linear regression with group A set as the reference. Sex and number of common albinism-associated alleles were used as covariates (Methods, Supplementary Fig.4). A p-value of $2.2 \times 10^{-16}$ was obtained for the model overall.

The number of individuals in the A, B, C, and D groups were as follows: 6,870; 5,976; 112; and 96.

**Supplementary Table 7**. Single-nucleotide variants used to calculate the genomic inflation factor lambda (λGC)

| variant coordinates (GRCh38) | variant dbSNP rsID | variant consequence (MANE Select v0.95 transcript) | name of associated gene (HGNC) | $\chi^2$ (Firth regression) | gnomAD v3.1.2 frequency in NFE [a] | CADD Phred score [b] | SpliceAI score |
|---|---|---|---|---|---|---|---|
| 12-88507004-A-T | rs183924903 | ENST00000644744.1: c.714+24T>A | *KITLG* | 0.099 | 0.001705 | 0.002 | 0 |
| 3-52404411-C-T | rs150454901 | ENST00000460680.6: c.1250+42G>A | *BAP1* | 0.976 | 0.003101 | 0.017 | 0 |
| 3-119490496-A-G | rs142663532 | ENST00000295588.9: c.798-55A>G | *POGLUT1* | 0.314 | 0.00369 | 0.041 | 0 |
| 16-84016604-G-A | rs77876966 | ENST00000299709.8: p.Leu359Leu | *SLC38A8* | 0.864 | 0.05285 | 0.075 | 0.02 |
| 5-33944723-G-A | rs150473213 | ENST00000296589.9: p.Val506Val | *SLC45A2* | 9.828 | 0.0002205 | 0.077 | 0 |
| 15-58646011-A-G | rs201948093 | ENST00000260408.8: c.735+44T>C | *ADAM10* | 0.18 | 0.003235 | 0.191 | 0 |
| 6-397261-G-A | rs34318727 | ENST00000380956.9: c.637+9G>A | *IRF4* | 16.21 | 0.07294 | 0.193 | 0 |
| 3-133953627-T-C | rs199782188 | ENST00000310926.11 : c.724+36A>G | *SLCO2A1* | 0.042 | 0.01306 | 0.222 | 0 |
| 1-154589737-T-G | rs115805812 | ENST00000368474.9: c.2668+20A>C | *ADAR* | 0.039 | 0.004735 | 0.247 | 0 |
| 10-98429811-C-A | rs11592273 | ENST00000361490.9: p.Gly283Trp | *HPS1* | 1.937 | 0.0758 | 0.337 | 0 |
| 15-58646039-G-A | rs138743329 | ENST00000260408.8: c.735+16C>T | *ADAM10* | 0.409 | 0.00178 | 0.385 | 0 |
| 3-69949177-C-G | rs181810413 | ENST00000352241.9: c.880+9C>G | *MITF* | 0.414 | 0.003381 | 0.553 | 0 |
| 15-55228733-T-C | rs368760836 | ENST00000336787.6: c.240-21A>G | *RAB27A* | 2.213 | 0.0001176 | 0.611 | 0 |
| 20-32230808-G-A | rs200896099 | ENST00000375749.8: c.736-11G>A | *POFUT1* | 0.143 | 0.001911 | 0.685 | 0.09 |
| 19-2114320-G-A | rs117387599 | ENST00000643116.3: c.2424-18C>T | *AP3D1* | 2.605 | 0.01143 | 0.698 | 0 |
| 15-27871136-T-C | rs41304383 | ENST00000354638.8: c.2244+18A>G | *OCA2* | 5.378 | 0.04011 | 0.831 | 0 |
| 4-54709427-C-T | rs72549293 | ENST00000288135.6: p.Tyr373Tyr | *KIT* | 0.983 | 0.0001029 | 0.894 | 0.03 |
| 12-52516840-G-A | rs144359915 | ENST00000252242.9: p.Asn412Asn | *KRT5* | 0.021 | 0.002307 | 0.94 | 0 |
| 9-21974641-C-G | rs45456595 | ENST00000304494.9: c.150+37G>C | *CDKN2A* | 0.177 | 0.003248 | 1.22 | 0.29 |
| 1-154597286-C-T | rs200651742 | ENST00000368474.9: c.1935-19G>A | *ADAR* | 0.069 | 0.00172 | 1.26 | 0 |
| 6-15524467-T-C | rs16876573 | ENST00000344537.10 : c.811+59A>G | *DTNBP1* | 0.472 | 0.04261 | 1.9 | 0 |
| 4-174492145-C-T | rs17060532 | ENST00000296522.11 : c.663-51G>A | *HPGD* | 0.016 | 0.04114 | 2.05 | 0 |
| 6-131851119-A-G | rs200746177 | ENST00000647893.1: c.431-23A>G | *ENPP1* | 0.375 | 0.001323 | 2.26 | 0 |

(continued)

| variant coordinates (GRCh38) | variant dbSNP rsID | variant consequence (MANE Select v0.95 transcript) | name of associated gene (HGNC) | $\chi^2$ (Firth regression) | gnomAD v3.1.2 frequency in NFE [a] | CADD Phred score [b] | SpliceAI score |
|---|---|---|---|---|---|---|---|
| 6-15533192-C-A | rs75380691 | ENST00000344537.10: c.667+48G>T | *DTNBP1* | 1.93 | 0.005808 | 2.69 | 0.04 |
| 12-88506969-A-G | rs41283110 | ENST00000644744.1: c.714+59T>C | *KITLG* | 4.972 | 0.007056 | 2.76 | 0 |
| 2-219212539-T-C | rs114354921 | ENST00000265316.9: c.1864-48A>G | *ABCB6* | 0.039 | 0.005657 | 3.11 | 0 |
| 15-52372211-G-A | rs11637651 | ENST00000399233.6: p.Ile910Ile | *MYO5A* | 0.892 | 0.06795 | 3.24 | 0 |
| 12-52519946-G-A | rs11549951 | ENST00000252242.9: p.Leu117Leu | *KRT5* | 0.359 | 0.07463 | 3.29 | 0 |
| 12-52517611-G-A | rs149467228 | ENST00000252242.9: p.Ala357Ala | *KRT5* | 1.656 | 0.007071 | 3.56 | 0.01 |
| 19-2113468-G-A | rs146083008 | ENST00000643116.3: c.2602-55C>T | *AP3D1* | 0.029 | 0.02771 | 3.83 | 0.06 |
| 3-149155185-G-A | rs34197730 | ENST00000296051.7: p.Thr493Thr | *HPS3* | 0.601 | 0.03576 | 4.28 | 0.13 |
| 12-57751138-C-G | rs3211614 | ENST00000257904.11: c.355-48G>C | *CDK4* | 1.646 | 0.002073 | 4.51 | 0.06 |
| 12-88516544-G-T | rs182948635 | ENST00000644744.1: c.364-54C>A | *KITLG* | 0.02 | 0.006951 | 4.53 | 0 |
| 2-237540465-G-T | rs61737681 | ENST00000264605.8: p.Ala408Ser | *MLPH* | <0.00001 | 0.03652 | 0.116 | 0 |
| 10-102065990-G-A | rs3737243 | ENST00000299238.7: p.Gly172Gly | *HPS6* | 6.705 | 0.1081 | 4.59 | 0 |

[a] gnomAD v3.1.2 frequency in NFE corresponds to minor allele frequency in the non-Finnish European subset of the Genome Aggregation Database (gnomAD) v3.1.2.

[b] CADD corresponds to Combined Annotation-Dependent Depletion, an integrative annotation tool for genetic variants. CADD provides a ranking rather than a prediction and the PHRED-scaled score ranges from 1 to 99 (with higher values indicating more deleterious cases). For the λGC analysis we wanted to select variants that are unlikely to affect protein function and we only chose changes with a CADD PHRED-scaled score less than 5.

It is noted that $\lambda_{median}$ was found to be 1.04. Given that this is close to the expected value of 1.00, it can be concluded that there is limited evidence of confounding by ancestry.

**Supplementary Table 8**. Case-control comparisons between different groups of genotypes linked to the *TYR*:c.1205G>A (p.Arg402Gln) and *OCA2*:c.1327G>A (p.Val443Ile) variants:

Genomics England 100,000 Genomes Project dataset analysis [a]

| genotype | | | | | | |
|---|---|---|---|---|---|---|
| ***TYR:*** **c.1205G>A (p.Arg402Gln) [rs1126809]** | ***OCA2*: c.1327G>A (p.Val443Ile) [rs74653330]** | **group** | **odds ratio** [b] | **95% confidence interval** | **unadjusted p-value** | **number of probands (cases / controls)** [a] |
| GG | GG | *A* | 1 | not applicable | | 12 / 10,369 |
| AG | GG | *B* | 2.6 | 1 – 7 | 0.05 | 15 / 8,011 |
| GG | AG | *C* | 2.5 | 0 – 19.3 | 0.57 | 0 / 158 |
| AG | AG | *D* | 14.8 | 1.5 – 71.9 | 0.026 | 1 / 128 |
| AA [c] | GG | *E* | 4.5 | 0.8 – 25.7 | 0.09 | 3 / 1,658 |
| AA [c] | AG | *F* | 41.2 | 0.3 – 520.1 | 0.10 | 0 / 26 |
| GG | AA | | these genotypes were not studied as individuals in these groups are considered to have a molecular diagnosis (*i.e.* to be “solved”) and therefore did not meet the inclusion criteria for analysis | | | |
| AG | AA | | | | | |
| AA | AA | | | | | |

[a] In contrast to the primary analysis presented in Fig.2, Supplementary Fig.1a and Supplementary Table 2, this secondary analysis focused only on individuals (both cases and controls) from the Genomics England 100,000 Genomes Project dataset. This study was conducted only on individuals who: (i) were projected to have European-like ancestries, (ii) were not found to have genotypes in keeping with a molecular diagnosis of albinism, (iii) carried no pathogenic or likely pathogenic variants in the *TYR* or *OCA2* gene (with the exception of the two studied changes *TYR*:c.1205G>A and *OCA2*:c.1327G>A). Ancestry was estimated using principal component analysis in all cases and controls.

[b] Firth logistic regression was used to generate odds ratios and unadjusted p-values; 35 probands with albinism and 20,304 unrelated controls were included in this analysis. Sex and the number of common albinism-associated alleles were used as covariates. The presented odds ratio corresponds to the probability of receiving a diagnosis of albinism. Group A was used as the reference group to which all other groups were compared to; the odds ratio for this group was therefore fixed at 1.

[c] As only individuals without a molecular diagnosis were considered, these groups include study subjects who were homozygous for *TYR*:c.1205G>A but did not carry the *TYR* c.-301= or the *TYR* c.575C>A variant in the homozygous state.

A plot of these results can be found in Supplementary Fig.1b

**Supplementary Table 9**. Unsolved – solved case comparisons between different groups of genotypes linked to the *TYR*:c.1205G>A (p.Arg402Gln) and *OCA2*:c.1327G>A (p.Val443Ile) variants:

University Hospital of Bordeaux albinism cohort [a]

| genotype | | | number of probands (unsolved cases / solved cases) [a] | p-value ($\chi^2$) [b] | odds ratio [c] | 95% confidence interval | p-value (Benjamini-Hochberg adjusted) |
|---|---|---|---|---|---|---|---|
| ***TYR:*** **c.1205G>A (p.Arg402Gln) [rs1126809]** | ***OCA2*****: c.1327G>A (p.Val443Ile) [rs74653330]** | **group** | | | | | |
| GG | GG | *A* | 91 / 255 | 0.02 | 1.6 | 1.15 – 2.27 | 0.021 |
| AG | GG | *B* | 56 / 242 | | 0.75 | 0.53 – 1.08 | 0.17 |
| GG | AG | *C* | 7 / 26 | | 1.03 | 0.42 – 2.26 | 0.934 |
| AG | AG | *D* | 9 / 12 | | 3 | 1.22 – 7.2 | 0.035 |

[a] In contrast to the primary and secondary analyses presented in Fig.2, Supplementary Fig.1 and Supplementary Tables 2 and 8, this analysis focused only on individuals from the University Hospital of Bordeaux albinism cohort. After excluding study subjects who did not report having European-like ancestries, the cohort was split into cases without and cases with a molecular diagnosis of albinism (i.e., “unsolved” and “solved”). Individuals without a molecular diagnosis who carried pathogenic or likely pathogenic variants in the *TYR* or *OCA2* gene (with the exception of the two studied changes *TYR*:c.1205G>A and *OCA2*:c.1327G>A) were then removed from the unsolved group. The patterns observed for genotype groups A-D were then inspected in the unsolved and solved cohorts.

[b] The patterns of variation for the categorical variable “unsolved to solved case ratio” were evaluated in genotype groups A-D using a $\chi^2$ test. The obtained p-value suggests that there are significant differences between the groups. Extensive pair-wise comparisons (post hoc tests) were not undertaken. However, individuals who carried genotype group D were compared to all other (*i.e.* non-group D) probands using a $\chi^2$ test; this revealed a p-value of 0.016.

[c] Firth logistic regression was used to generate odds ratios and unadjusted p-values; 173 unsolved and 628 solved cases were included in this analysis. Sex and the number of common albinism-associated alleles were used as covariates. Since (i) only affected individuals were evaluated and (ii) a reference group was not assigned, the odds ratios presented in this table should not be compared or interpreted in a similar fashion to those presented in Supplementary Tables 2, 3 and 8.

**Supplementary Table 10**. Albinism case definition in the different cohorts used in this study

| cohort | overall number of cases | approach/criteria |
|---|---|---|
| University Hospital of Bordeaux Molecular Genetics Laboratory cohort | 1,015 | • identified through the database of the University Hospital of Bordeaux Molecular Genetics Laboratory, France.<br>• independent assessment of available phenotypic data was performed by two clinicians with experience in albinism diagnostics; only cases with a consensus clinical impression of albinism were included [a]. |
| Genomics England 100,000 Genomes Project cohort | 105 | • enrolled across multiple clinical specialties in designated Genomic Medicine Centres (GMC) within the English National Health Service (NHS); standardised baseline clinical data were recorded with the use of Human Phenotype Ontology (HPO) terms guided by disease-specific data models.<br>• primary clinical data were inspected; individuals with one of the following codes were included in the relevant case group:<br>o the ICD-10 term “Albinism” [E70.3]<br>o the HPO term “Albinism” [HP:0001022]<br>o the HPO term “Partial albinism” [HP:0007443]<br>o the HPO term “Ocular albinism” [HP:0001107]. |
| UK Biobank cohort | 39 | • recruited in dedicated UK Biobank assessment centres; the baseline visit included a touchscreen questionnaire, physical measures and biological sampling; crucially, a range of data from other national datasets were incorporated including primary care, screening programmes, and disease-specific registries.<br>• a summary of the distinct ICD-10 diagnosis codes that each participant has had recorded across all their hospital inpatient records (in either the primary or secondary position) was available in data field 41270.<br>• primary care data recorded by health professionals working at general practices was available in resource 591 (category 3000).<br>• UK Biobank participants with the ICD-10 term “Albinism” [E70.3] in data field 41270 or resource 591 were included in the relevant case group. |

[a] The principle that underlies this process is that the diagnosis of albinism, like many other diagnoses in medicine, is based on a gestalt, a constellation of observations and findings. There is no single sign or symptom that is perfectly diagnostic and clarity can only be gained when different pieces of available information (clinical, genetic, imaging, electrophysiological etc) are jointly inspected and combined using Bayesian reasoning.

**ACKNOWLEDGEMENTS & CONSORTIA**

We are very grateful to individuals and families who are included in the studied cohorts for their participation. We are also grateful to the clinical, research and support staff who helped with recruitment and to the teams who are managing these cohorts and the associated resources.

We acknowledge the contribution of the Genomics England Research Consortium. The members of this consortium are: John C. Ambrose, Prabhu Arumugam, Roel Bevers, Marta Bleda, Freya Boardman-Pretty, Christopher R. Boustred, Helen Brittain, Mark J. Caulfield, Georgia C. Chan, Greg Elgar, Tom Fowler, Adam Giess, Angela Hamblin, Shirley Henderson, Tim J. P. Hubbard, Rob Jackson, Louise J. Jones, Dalia Kasperaviciute, Melis Kayikci, Athanasios Kousathanas, Lea Lahnstein, Sarah E. A. Leigh, Ivonne U. S. Leong, Javier F. Lopez, Fiona Maleady-Crowe, Meriel McEntagart, Federico Minneci, Loukas Moutsianas, Michael Mueller, Nirupa Murugaesu, Anna C. Need, Peter O'Donovan, Chris A. Odhams, Christine Patch, Mariana Buongermino Pereira, Daniel Perez-Gil, John Pullinger, Tahrima Rahim, Augusto Rendon, Tim Rogers, Kevin Savage, Kushmita Sawant, Richard H. Scott, Afshan Siddiq, Alexander Sieghart, Samuel C. Smith, Alona Sosinsky, Alexander Stuckey, Mélanie Tanguy, Ana Lisa Taylor Tavares, Ellen R. A. Thomas, Simon R. Thompson, Arianna Tucci, Matthew J. Welland, Eleanor Williams, Katarzyna Witkowska, Suzanne M. Wood

We also acknowledge the contribution of the UK Biobank Eye and Vision Consortium. Members of this consortium are: Naomi Allen, Tariq Aslam, Denize Atan, Sarah Barman, Jenny Barrett, Paul Bishop, Graeme Black, Tasanee Braithwaite, Roxana Carare, Usha Chakravarthy, Michelle Chan, Sharon Chua, Alexander Day, Parul Desai, Bal Dhillon, Andrew Dick, Alexander Doney, Cathy Egan, Sarah Ennis, Paul Foster, Marcus Fruttiger, John Gallacher, David Garway-Heath, Jane Gibson, Jeremy Guggenheim, Chris Hammond, Alison Hardcastle, Simon Harding, Ruth Hogg, Pirro Hysi, Pearse Keane, Peng Tee Khaw, Anthony Khawaja, Gerassimos Lascaratos, Thomas Littlejohns, Andrew Lotery, Robert Luben, Phil Luthert, Tom Macgillivray, Sarah Mackie, Savita Madhusudhan, Bernadette Mcguinness, Gareth Mckay, Martin Mckibbin, Tony Moore, James Morgan, Eoin O'Sullivan, Richard Oram, Chris Owen, Praveen Patel, Euan Paterson, Tunde Peto, Axel Petzold, Nikolas Pontikos, Jugnoo Rahi, Alicja Rudnicka, Naveed Sattar, Jay Self, Panagiotis Sergouniotis, Sobha Sivaprasad, David Steel, Irene Stratton, Nicholas Strouthidis, Cathie Sudlow, Zihan Sun, Robyn Tapp, Dhanes Thomas, Emanuele Trucco, Adnan Tufail, Ananth Viswanathan, Veronique Vitart, Mike Weedon, Cathy Williams, Katie Williams, Jayne Woodside, Max Yates, Jennifer Yip, Yalin Zheng.

This research was conducted using the UK Biobank Resource under projects 53144 and 49978.
